# Postprandial amino acid profiles and gastric behavior after consumption of milk and plant-based alternatives: a randomized cross-over study in healthy males

**DOI:** 10.64898/2026.09.23.26363226

**Authors:** Louise M. Leenders, Silvia Casagranda, Michiel G.J. Balvers, Gerben de Gier, Tim T. Lambers, Paul A.M. Smeets

## Abstract

**Background:** There is a growing availability of plant-based alternatives for bovine milk. However, plant-based drinks differ e.g. in protein quantity, protein quality and carbohydrate composition. Knowledge on the impact this has on digestion-related outcomes such as postprandial amino acid (AA) availability and gastric behavior is still limited. Additionally, data on milk alternatives with higher overall protein concentration are lacking.

**Objective:** We aimed to compare the differences in postprandial AA profiles, glycemic response, and gastric behavior between bovine milk and plant-based drinks with or without added protein using representative commercial drinks.

**Methods:** 12 healthy males participated in a randomized crossover study with three treatments. After an overnight fast, participants drank 750 ml of semi-skimmed milk (Milk), a standard oat-based drink (OBD) or an oat-based drink enriched with pea protein (OBD+Pea). Blood samples were taken for measurement of plasma AA, glucose, insulin and FGF21 over 5 hours. Gastric content changes were assessed using magnetic resonance imaging scans for Milk and OBD+Pea over 2 hours.

**Results:** Compared with Milk, both oat-based drinks elicited lower 5-h total AA iAUC (OBD-169706; OBD+pea -139552 µM·min; both P<0.001) and EAA iAUC (OBD -91748; OBD+pea-78199; both P<0.001), with no differences between oat-based drinks. Gastric content volume was lower for OBD+Pea versus Milk from 60-120 min (range -48 – -84 ml, all P<0.001). Compared to Milk, OBD and OBD+Pea increased glucose more from 15-45 min (range 0.86–1.73 mmol/l, all P<0.001) and insulin from 30-60 min (1.84–2.14-fold, all P<0.001).

**Conclusions:** Protein enrichment in plant-based milk alternatives can be a valuable strategy to increase protein content and improve amino acid composition. However, such products still show faster gastric emptying, lower postprandial essential amino acid and higher glycemic and insulinemic responses compared to bovine milk because of their different carbohydrate and protein content and quality.

## Introduction

There is growing interest in plant-based diets among consumers, because of their potential health benefits and positive environmental impact [1,2]. In line with this, the availability of plant-based food products is rapidly growing. This includes alternatives for cow’s and other animal’s milk. Accordingly, the availability of plant-based drinks in supermarkets has sharply increased. However, this rise in plant-based alternatives for milk can be concerning from a health perspective because many are not as nutritious as animal milk [3], and from that perspective should not be used as a one-on-one substitute, especially in specific populations such as infants, children, diseased and elderly individuals [4,5]. However, consumers may not be aware of this because these products are often sold as an alternative for milk [6]. Plant-based drinks contain lower amounts of protein and other important nutrients, such as calcium and vitamin B_12_. To compare, bovine milk contains about 3.5 g of protein per 100 ml and most plant-based alternatives in the Netherlands contain 0.1 – 0.9 g of protein per 100 ml, with the exception of soy-based drinks which contain around 3.0 – 3.5 g of protein per 100 ml [6,7]. In addition to their generally lower protein content, plant-based alternatives have lower protein quality as reflected by their Digestible Indispensable Amino Acid Score (DIAAS), which provides an indication of protein quality according to the ileal digestibility and adequacy of individual indispensable amino acids (AAs) relative to human requirements [8]. Proteins with a DIAAS ≥100 are considered “excellent” sources, while scores between 75–99 indicate “good” quality. For bovine milk, the DIAAS is approximately 114, whereas plant-based protein scores generally range from 29-89. Among plant protein sources, only soy, pea protein concentrate and chickpea have scores indicating a “good quality protein”, with scores of 89, 82, and 83, respectively [9]. However, these scores do not account for the protein concentration, which is generally lower in plant-based drinks compared to bovine milk.

Differences in protein quantity and quality between bovine milk and plant-based alternatives are also important because of their potential influence on gastric digestion, which can subsequently impact postprandial amino acid responses. For instance, the casein fraction of bovine milk can coagulate during gastric digestion *in vivo* [10,11]. This can alter gastric emptying, with the liquid fraction emptying faster than semi-solid coagula, which leads to later appearance of AAs from the coagulum in the circulation [12,13]. Evidence for gastric coagulation of non-milk proteins is limited, with pea protein isolate reported to form only tiny particles with no apparent effect on gastric emptying [14]. In how far proteins coagulate in the stomach not only depends on acidity and protease activity but also on the preceding processing of the protein product, in particular heat treatment [15–17].

Differences in protein quality between bovine milk and plant-based alternatives can also impact post-prandial glucose and insulin responses, as can differences in their carbohydrate composition; for bovine milk lactose is the predominant carbohydrate while plant-based milk alternatives contain a variety of other carbohydrates [18]. Although the higher carbohydrate content of plant-based drinks is the primary determinant of blood glucose and insulin responses, ingested protein also has an insulinotropic effect, particularly when it is rich in branched-chain amino acids (BCAAs), which are generally more abundant in animal products [19–21]. Moreover, lactose has a lower glycemic index than carbohydrates generally present in plant-based drinks [22,23].

Differences in protein content and quality can also alter fibroblast growth factor 21 (FGF21) secretion. Circulating FGF21 concentrations increase in response to protein restriction [24–26], especially with methionine restriction [27,28], driving adaptations in macronutrient preference that favor protein intake. However, effects of protein quality on circulating FGF21, especially in the context of real world products, are not known. Next to ensuring the ingestion of a sufficient amount of protein it is also important to ensure adequate protein quality. This is particularly relevant for specific populations. For example, it is known that elderly have difficulties consuming sufficient protein [29]. Furthermore, for children, dairy can be an important contributor to meeting nutritional requirements for healthy growth and bone development, which may not be met by plant-based drinks [30,31]. Also, in disease and disease-related malnutrition, protein quality is important to maintain muscle mass and promote recovery [32].

Due to differences in protein content and quality, it is important to compare the digestion of milk with that of plant-based alternatives. In addition, it is important to explore the effect of increasing both protein content and protein quality by enriching plant-based drinks with other plant proteins. Blends of legume and grain-derived protein sources may, to a certain extent, complement essential amino acid (EAA) limitations of the individual sources. For example, it has been suggested that oat drinks could be fortified with pea protein to improve the EAA composition [7].

Accordingly, this study aimed to compare digestion-related responses and gastric behavior following ingestion of semi-skimmed milk with those elicited by a commercially available oat-based drink and a similar oat-based drink enriched with pea protein. In addition, intragastric behavior was measured with magnetic resonance imaging (MRI) for milk and the protein-enriched oat-based drink to determine potential differences in gastric protein coagulation and gastric emptying patterns. This design allows a quantitative *in vivo* comparison of the postprandial AA kinetics of bovine milk and real-world plant-based alternatives, mimicking the nutritional impact of replacing milk with a plant-based drink.

## Participants and Methods

### Study design

This study was a double-blind randomized crossover trial in which healthy participants underwent blood sampling at baseline and after consumption of semi-skimmed milk, an oat-based drink or an oat-based drink with added pea protein. For the milk and the enriched oat-based drink abdominal MRI scans were performed as well (**Figure 1**).

**Figure 1.**
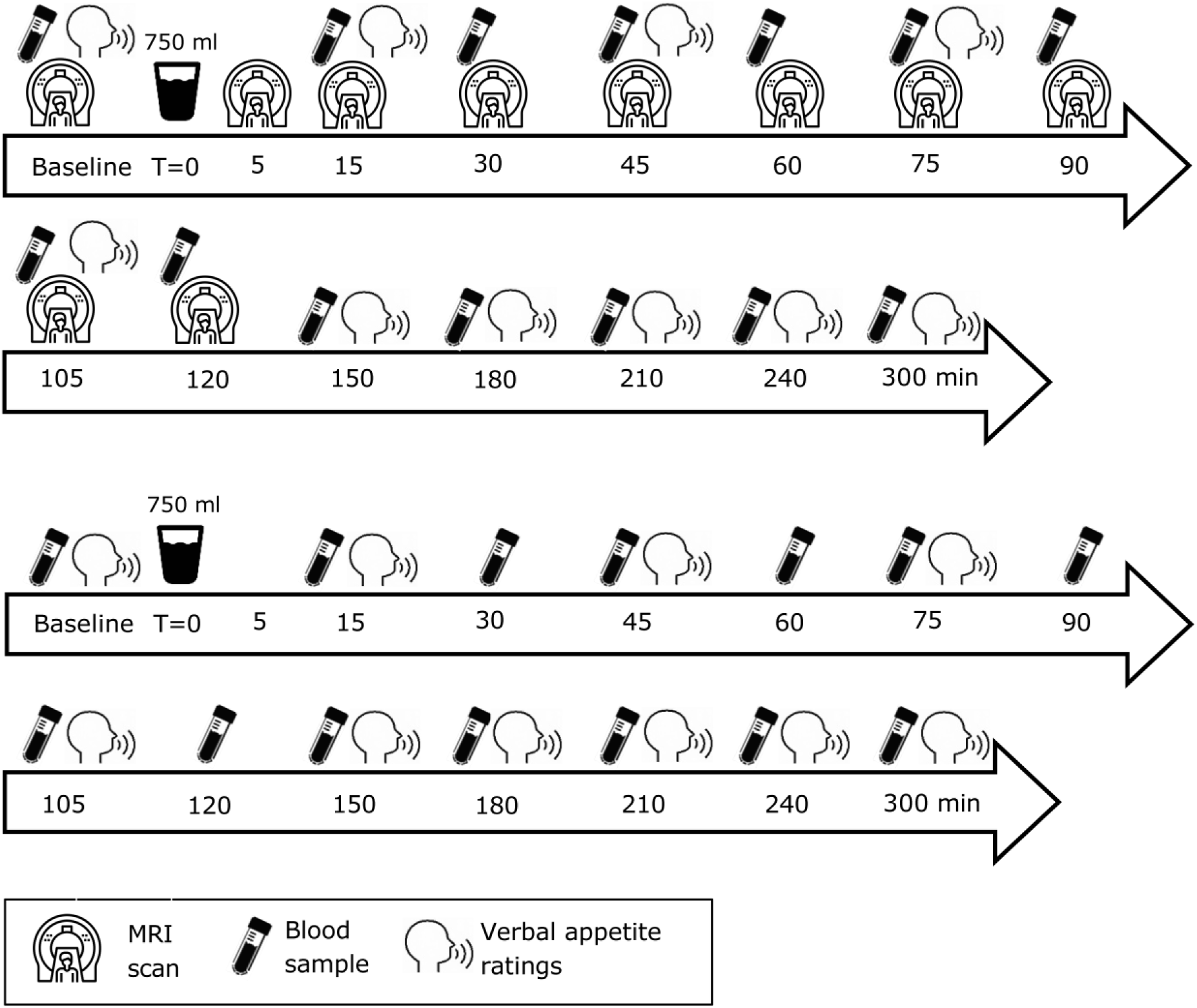
Overview of the two types of test days. Top: test days with MRI (Milk, OBD+Pea. Bottom: Test days without MRI (OBD). T=0 min is the start of consumption.

The primary outcome was the change in postprandial plasma amino acid profile over time. Secondary outcomes were gastric behavior (gastric content volume over time, coagulation) and plasma glucose and insulin. Other reported outcomes were subjective ratings regarding wellbeing and appetite. FGF21 [25] was assessed as an exploratory outcome.

The study procedures were approved by an accredited Medical Ethical Committee (METC Oost-Nederland) in accordance with the Helsinki Declaration of 1975 as revised in 2013. The study was registered with clinicaltrials.gov under number NCT06272331 in February 2022. All participants signed informed consent.

### Sample size estimation

Due to the exploratory nature of the study, there were no literature examples that were directly comparable to our products and study design, nor did we aim to do formal hypothesis testing. To assess our ability to detect relevant differences between the treatments data from Liu et al. [33] who compared amino acid profiles between dairy and vegetable proteins in healthy elderly were used. The sample size calculation was performed for the difference in peak postprandial AA concentration and the difference in the area under the curve of plasma amino acid concentration over time. A two-sided paired test was deemed appropriate, with a significance level of 0.05 and a power of 90%. The power calculation showed that for a peak within-subject difference of 500 μmol/l with an SD of 350 μmol/l, a total of 7 complete datasets would be needed. For the area under the curve a difference of 43000 μmol/l*min, combined with a SD of 40000 μmol/l*min, would require 11 complete datasets. Thus, a meaningful difference in the AA response could be detected with a sample size of 12 participants, accounting for possible drop-out.

### Participants

We recruited apparently healthy (self-reported) men aged 18–45 years with a BMI between 18.5 and 25 kg/m^2^. Exclusion criteria were: Blood hemoglobin concentration below 8.5 mmol/l (as measured with a finger-prick test); Having a gastric disorder or regular gastric complaints, such as heart burn; Use of medication which alters the normal functioning of the stomach, such as medical drugs that influence the gastrointestinal tract’s normal function (e.g. protein pump inhibitors, antacids, anti-depressants, as judged by a medical professional) or the microbiota (e.g. antibiotics use within one month prior to the pre-study screening day); Following a vegan diet; Allergy or intolerance for cow milk, lactose or gluten (self-reported); Smoking (>2 cigarettes/week); Alcohol consumption of more than 14 glasses/week; Having a gastric disorder or regular gastric complaints such as heart burn (more than once per week); Having a chronic illness that could affect food digestion and nutrient absorption including but not limited to kidney disease, thyroid disease and diabetes mellitus (self-reported); Having a contra-indication to MRI scanning (including but not limited to pacemakers and defibrillators, ferromagnetic implants, and claustrophobia).

Participants were recruited between January and April 2024 via digital advertisements (e-mail and social media) and invited for an information session where they could ask questions. Those that appeared eligible and were willing to participate were invited for a screening session. During this session they signed informed consent, filled in the inclusion questionnaire, had their hemoglobin concentrations checked (finger prick). To avoid dropouts, participants tasted a glass of one of the oat-based drinks after which they rated their liking on a 9-point Likert scale and confirmed verbally their ability to consume 750 ml on the test days. N=12 men (age 27 ± 7.4 years; BMI 22.8 ± 2.3 kg/m^2^) were included in the study and completed three test sessions between March and July 2024 (see **Supplementary Figure 1**).

### Treatments

The three treatments consisted of 750 ml semi-skimmed UHT-treated bovine milk (Milk), a regular oat-based drink (OBD), and an oat-based drink with 1 g/100 ml pea protein added (OBD+Pea, FrieslandCampina) obtained from the local supermarket. Their detailed composition can be found in **Supplementary Table 1**. The OBD+Pea drink contained a similar amount of energy and fat as the milk but less protein (12.8 versus 27 g). They were consumed at room temperature from an opaque cup and participants were instructed to finish within 5 min, which they all did. Participants were not informed about the type of drink they received. Treatment order was randomized by a researcher using simple randomization. Allocation sequences were generated in Microsoft Excel using its random number generation function.

### Study procedures

Study procedures are summarized in Figure 1. Subjects were instructed to consume the same evening meal before each visit, to refrain from alcohol consumption that evening, and to fast from 10 p.m. onwards (no food or drinks except water). Drinking water was allowed up to 90 min prior to their visit. Upon arrival at Hospital Gelderse Vallei (Ede, The Netherlands), a cannula was placed in an antecubital vein. Subsequently, baseline blood samples were taken, MRI scans were performed, and verbal ratings were obtained regarding wellbeing (nausea, stomach complaints) and appetite (hunger, fullness, thirst). Ratings were given on a 100-unit scale with 0 representing “not at all” and 100 “very much” [34]. After this, participants were instructed to drink 750 ml of one of the drinks through a straw in 5 min while seated on the scanner bed. All participants successfully managed to consume the drink within 5 min. Immediately after finishing the drink, the participant was returned to a supine position on the scanner bed. Postprandial gastric MRI scans were performed at time points t=5, 15, 30, 45, 60, 75, 90, 105, and 120 min post-consumption and blood samples were taken at t=15, 30, 45, 60, 75, 90, 105, 120 min. At t=150, 180, 210, 240, and 300 min participants sat upright in another room for blood collection. After each scan, participants rated their appetite and well-being at t=15, 45, 75, 105, 150, 180, 210, 240, and 300 min.

The OBD test session (blood sampling, but without MRI) was conducted on the Wageningen University campus (Figure 1 lower part). To mimic the MRI conditions of the other two test sessions, participants remained in a supine position on a bed for the first two hours. After this, they were seated in an upright position.

### Blood sample collection and analysis

Blood samples were drawn from the cannula into EDTA, sodium-fluoride and lithium heparin tubes. EDTA tubes were put on ice and centrifuged at 1300 g for 10 min at 4 °C within one hour of collection. Sodium-fluoride and lithium heparin tubes were kept at room temperature and centrifuged at 3000 *g* for 8 min at 22 °C, to obtain blood plasma. Blood plasma aliquots were stored at -80 °C until they were analyzed.

Amino acids: Detailed AA analytical procedures are provided in the Supplementary Materials. Briefly, EDTA plasma amino acid concentrations were quantified using UPLC-MS/MS based on a validated HILIC method [35]. Internal standards were added to sample aliquots and after protein precipitation the supernatant was injected into an Acquity H-Class Plus UPLC coupled to a Xevo TQ-S micro tandem mass spectrometer (Waters). Amino acids were separated using an Acquity UPLC BEH amide column (2.1 x 100 mm, 1.7 µm) and detected in positive electrospray ionization mode using multiple reaction monitoring and quantified from internal standard-corrected calibration curves (1/x² weighting). Inter-batch coefficients of variation were generally <15% for analytes with corresponding internal standards.

Glucose: To determine glucose concentration, the sodium fluoride plasma samples were processed using the Atellica CH Glucose Hexokinase_3 (GluH_3) assay kit and quantified using the Atellica CH analyzer (Siemens Healthineers). The detection range was 0.2 to 39 mmol/l, and the maximum intra-assay coefficient of variation was 4.5%. There were no glucose concentrations outside the detection range.

Insulin: The lithium heparin plasma samples were processed, and insulin was quantified using a solid-phase, enzyme-labeled chemiluminescent immunometric assay (IMMULITE 1000 Insulin) and an Atellica IM Analyzer (Siemens Healthineers). The detection range was 2 to 300 μIU/L, and inter-assay coefficients of variation ranged from 3.3% to 19.1%. Values below the detection limit were set at 1 μIU/l (lower limit/2) as a biologically plausible low value for healthy normal-weight young adults. There were no insulin concentrations above the detection limit. The Atellica analyzers are validated for single-replicate clinical measurements; no duplicates were analyzed. Because 89 of 503 concentrations (17.7%) were below the detection limit, the data were log-transformed before analysis.

FGF21: EDTA plasma concentrations were determined using an enzyme-linked immunosorbent assay (ELISA) method (Quantikine® ELISA, catalog number DF2100, R&D Systems) according to the manufacturer’s instructions. The detection range was 31.3 – 2000 pg/ml. 10% of the samples were analyzed in duplo. Coefficients of variation for those ranged from 0.1% to 10.2% (median 0.9, IQR = 1.2).

### MRI scanning

Participants were scanned head-first in a supine position with the use of a 3 Tesla Philips Ingenia Elition X MRI scanner (Philips, Eindhoven, The Netherlands) equipped with a dStream torso coil. Stomach scan (18 s): a 2-D Turbo Spin Echo sequence (37 4-mm slices, 1.4 mm gap, acquired in-plane voxel size 1.8 x 1.8 mm, reconstructed to 1 × 1 mm, repetition time 550 ms, echo time 80 ms, flip angle 90°, field of view 400 x 336 x 210 mm, SENSE factor anterior-posterior 2.5) was used with breath hold command on expiration to fixate the position of the diaphragm and the stomach.

### Stomach scan image analysis

Total gastric content volume was determined for each stomach scan with the help of an in-house software tool that performs a deep learning-based segmentation of the stomach contents based on an nnU-Net framework [36]. The resulting mask images were checked, and where necessary manually corrected, with the use of the ITK-SNAP program [37] (http://www.itksnap.org/). An example of a stomach scan time series for the two treatments is shown in **Supplementary Figure 2**. To assess potential changes in gastric coagulation two approaches were used; 1) Thresholding: To quantify the (relative) volume of liquid and more solid stomach contents the number of lighter (more liquid) and darker (more solid) voxels was calculated by determining an intensity threshold with the use of Otsu’s method [38] in Matlab (version R2023a, *multitresh* function), an approach previously used for MRI images of milk digestion [11,12,39,40]. 2) Image texture analysis: we calculated image texture features that quantify different aspects of spatial grey-level patterns in an image [41] for the stomach contents using LIFEx (version 7.6.8, Institut national de la santé et de la recherche médicale, France) [42]. This approach was previously applied in human MRI research on casein coagulation [10,43]. Specifically, we calculated Joint variance, Coarseness, Contrast, and Busyness. The Grey-Level Co-occurrence Matrix (GLCM) approach was used for Joint variance which reflects the variance of grey-level pairs around the mean intensity pair. Lower values mean that most voxel pairs have similar intensities, i.e., that image texture is more homogeneous. Neighborhood Grey Tone Difference Matrix (NGTDM) compares the grey level of each voxel with the average grey level of its local neighborhood and was used to calculate Contrast (local intensity variations), Coarseness (the spatial scale of texture patterns, with higher values indicating larger homogeneous regions), and Busyness (spatial frequency of intensity changes). The number of grey levels for texture metric calculation was set at 64, intensity rescaling at relative (ROI: min/max), and dimension processing at 3D. On each postprandial time point, texture metrics were calculated per slice for the stomach content. In the context of this study, changes in image texture metrics were interpreted as potentially reflecting changes in the degree of coagulation.

### Statistical analysis

Statistical analyses were performed using RStudio (Version 2026.04.0, Posit Software) and the R (R Core Team) software packages dplyr (1.2.1), pracma (2.4.6), nlme (3.1.169) and emmeans (2.0.3) on data from the 12 participants that completed three test sessions. Missing values were not imputed. For the amino acid analysis total AA concentration and essential AA (EAA) concentration were calculated by adding up all relevant individual amino acids. Subsequently, 5-h iAUC was calculated for all AA variables.

AA iAUC’s were analyzed using linear mixed models (lme), with treatment as a fixed factor, baseline concentration as a covariate and participant as a random factor (random intercept). Tukey-corrected post-hoc t-tests were used to compare estimated marginal means between the treatments.

Postprandial amino acid, glucose, insulin, gastric content volume and subjective ratings were analyzed using linear mixed models (lme), with the treatment*time interaction as a fixed factor, baseline values as a covariate and participant as a random factor (random intercept). For the image texture metrics the same model was used, but without a baseline covariate. Tukey-corrected post-hoc t-tests were used to compare estimated marginal means for the different treatments and time points. Insulin concentrations were natural log-transformed prior to analysis to improve model assumptions. Results are presented as differences in estimated marginal means with 95% confidence intervals (CIs). For insulin, model estimates were back-transformed and presented as ratios of geometric means with 95% CIs. Statistical significance was defined as a two-sided P-value <0.05.

## Results

### Amino acids

Total AA and EAA curves are shown in **Figure 2**. IAUC estimates and comparisons are detailed in **Supplementary table 4**. The 5-h iAUC for total AA concentrations was greater after milk compared with both OBD and OBD+Pea (both P<0.001). There was no iAUC difference between OBD and OBD+Pea (P=0.332). In line with this, total postprandial AA concentrations differed significantly by treatment over time (P<0.001). Milk elicited the highest amino acid response throughout most of the postprandial period, with concentrations peaking at 30–75 min (estimated marginal means 4425–4633 µmol/l) and remaining significantly greater than both OBD and OBD+Pea from 15 to 240 min (all P<0.002), except versus OBD+Pea at 240 min (P=0.062). OBD+Pea resulted in intermediate amino acid concentrations and those were significantly higher than for OBD from 15 to 105 min (all P<0.017), with the largest difference observed at 45 min (4052 versus 3450 µmol/l, *P*<0.0001). Thereafter, differences between the two plant-based beverages were no longer significant (120–300 min, all P*>*0.25). By 300 min, amino acid concentrations had declined toward baseline and no differences remained between treatments (all P>0.12).

**Figure 2.**
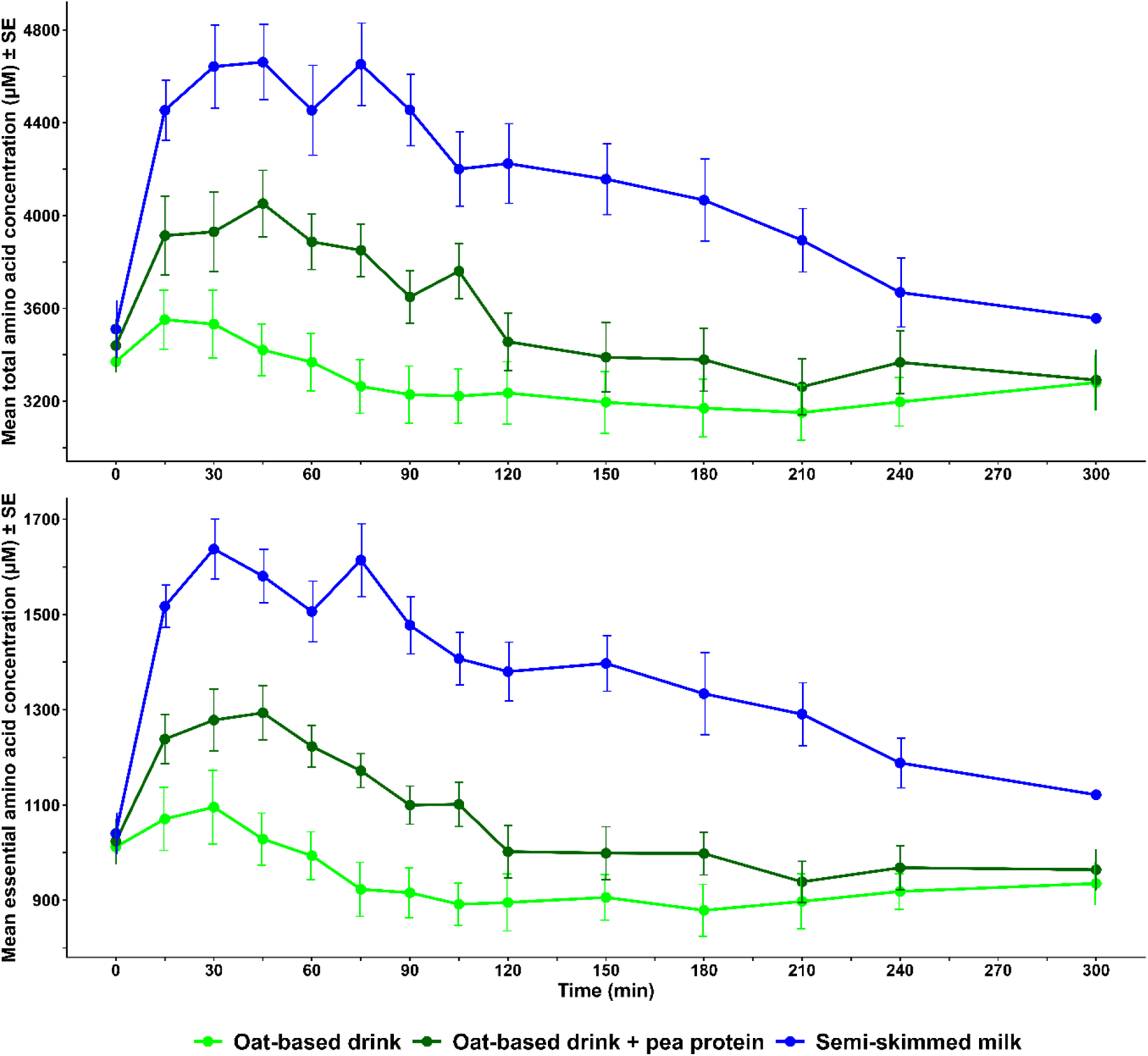
Mean ± SE total AA and EAA concentration over time for the three drinks. Postprandial concentrations were analyzed using linear mixed models with the treatment*time interaction as a fixed factor and baseline concentrations as a covariate.

Similarly, the 5-h iAUC for EAA was higher after Milk than after OBD or OBD+Pea (both P<0.001), whereas there was no difference between the two oat-based drinks (P=0.277). Similarly, postprandial EAA concentrations differed significantly among treatments throughout most of the postprandial period (interaction P<0.001). Milk elicited the highest EAA response, with concentrations peaking at 30–75 min (estimated marginal mean 1500–1631 µmol/l) and remaining significantly greater than both OBD and OBD+Pea from 15 to 300 min (all timepoints P<0.011). OBD+Pea produced intermediate EAA concentrations and that were significantly higher than for OBD from 15 to 105 min (all P≤0.005), with the largest difference observed at 45 min (1294 vs 1034 µmol/l, P<0.0001). From 120 min onwards, EAA concentrations no longer differed significantly between OBD and OBD+Pea (all P≥0.070). Although EAA concentrations declined after their early postprandial peak for all treatments, Milk maintained substantially higher concentrations throughout the 5-h postprandial period, whereas OBD and OBD+Pea converged from 120 min onwards. Detailed results are presented in Supplementary table 2 and 3. Curves for individual amino acids and iAUC differences between the drinks are presented in **Supplementary figures 3-5** and **Supplementary table 4**.

### Glucose and insulin

Curves are shown in **Figure 3**. Detailed results are presented in **Supplementary Table 5** and **6**. For glucose there was a treatment by time interaction such that glucose was higher for both oat-based drinks compared to Milk at T=15, 30 and 45 min (all P<0.001). At 15 min, glucose concentrations were higher for OBD and OBD+Pea than for Milk by 0.86 (95% CI (0.32, 1.40), P<0.001) and 0.95 mmol/L (95% CI (0.41, 1.49), P<0.001), respectively. Differences were greatest at 30 min, with glucose concentrations exceeding those for Milk by 1.68 (95% CI (1.13, 2.22), P<0.001) for OBD and 1.73 mmol/L (95% CI (1.19, 2.27), P<0.001) for OBD+Pea. At 45 min, glucose concentrations remained higher for OBD and OBD+Pea than for Milk by 1.22 (95% CI (0.68, 1.76), P<0.001) and 1.02 mmol/L (95% CI (0.48, 1.55), P<0.001), respectively. No differences were observed between OBD and OBD+Pea at any time point (all P≥0.65), and there were no treatment differences at later postprandial time points.

**Figure 3.**
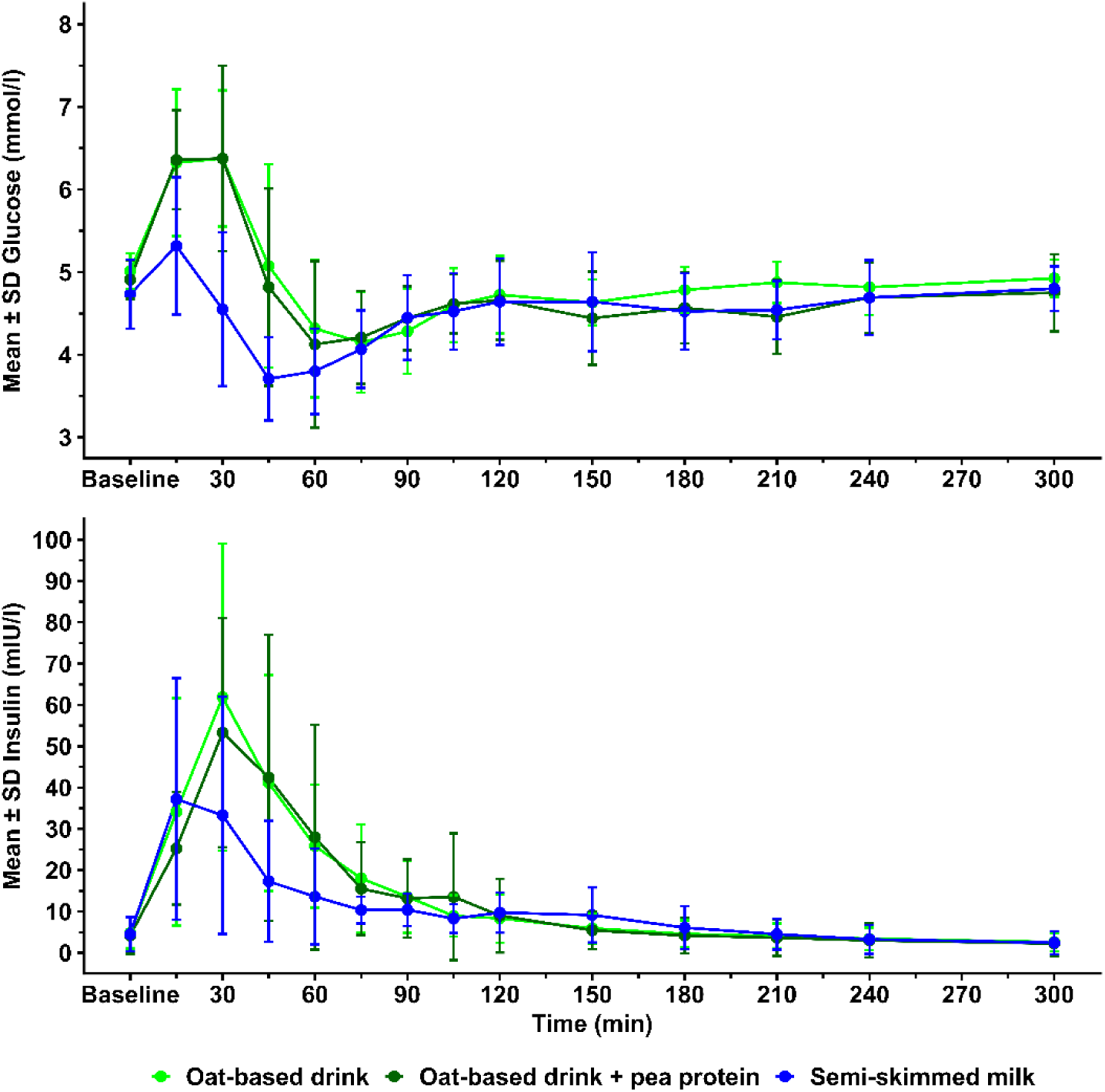
Mean ± SD glucose (top) and insulin (bottom) concentration over time for the three drinks. Postprandial concentrations were analyzed using linear mixed models with the treatment*time interaction as a fixed factor and baseline concentrations as a covariate.

For insulin there was also a treatment by time interaction (P<0.001). Compared with Milk, both OBD and OBD+Pea elicited significantly higher insulin concentrations from 30 until 60 min. At 30 min, insulin concentrations were 1.89-fold higher for OBD (95% CI (1.15, 3.09), P=0.007) and 1.84-fold higher for OBD+Pea (95% CI (1.12, 3.00), P=0.011) than for Milk. At 45 min, concentrations were 2.34-fold (95% CI (1.43, 3.83), P<0.001) and 2.54-fold (95% CI (1.55, 4.15), P<0.001) higher, respectively, and at 60 min they remained elevated 2.14-fold (95% CI (1.31, 3.50), P<0.001) and 2.09-fold (95% CI (1.28, 3.43), P=0.001), respectively. There were no differences in insulin concentration between OBD and OBD+Pea at any postprandial time point (all P>0.82).

### Gastric behavior

Changes in the gastric contents over time for OBD+Pea and Milk are illustrated in **Supplementary Figure 2.**

Gastric content volume (**Figure 4**): There was a significant treatment by time interaction (P=0.007). Postprandial volumes did not differ between OBD+Pea and Milk from 5 until 45 min (all P≥0.068), although there was a trend toward a lower gastric volume for OBD+Pea at 45 min (434 versus 466 ml, estimated marginal mean difference: −32 ml; 95% CI (−65, 2), P=0.068). From 60 min onward, gastric content volume was significantly lower following OBD+Pea than Milk, with mean treatment differences of −48 ml (95% CI (−82, −15), P=0.006) at 60 min, −56 ml (95% CI (−90, −22), P=0.001) at 75 min, −67 ml (95% CI (−101, −33), P<0.001) at 90 min, −84 mL (95% CI (−118, −50, P<0.001) at 105 min, and −68 ml (95% CI (−102, −34), P<0.001) at 120 min, indicating faster gastric emptying following OBD+Pea compared with Milk.

**Figure 4.**
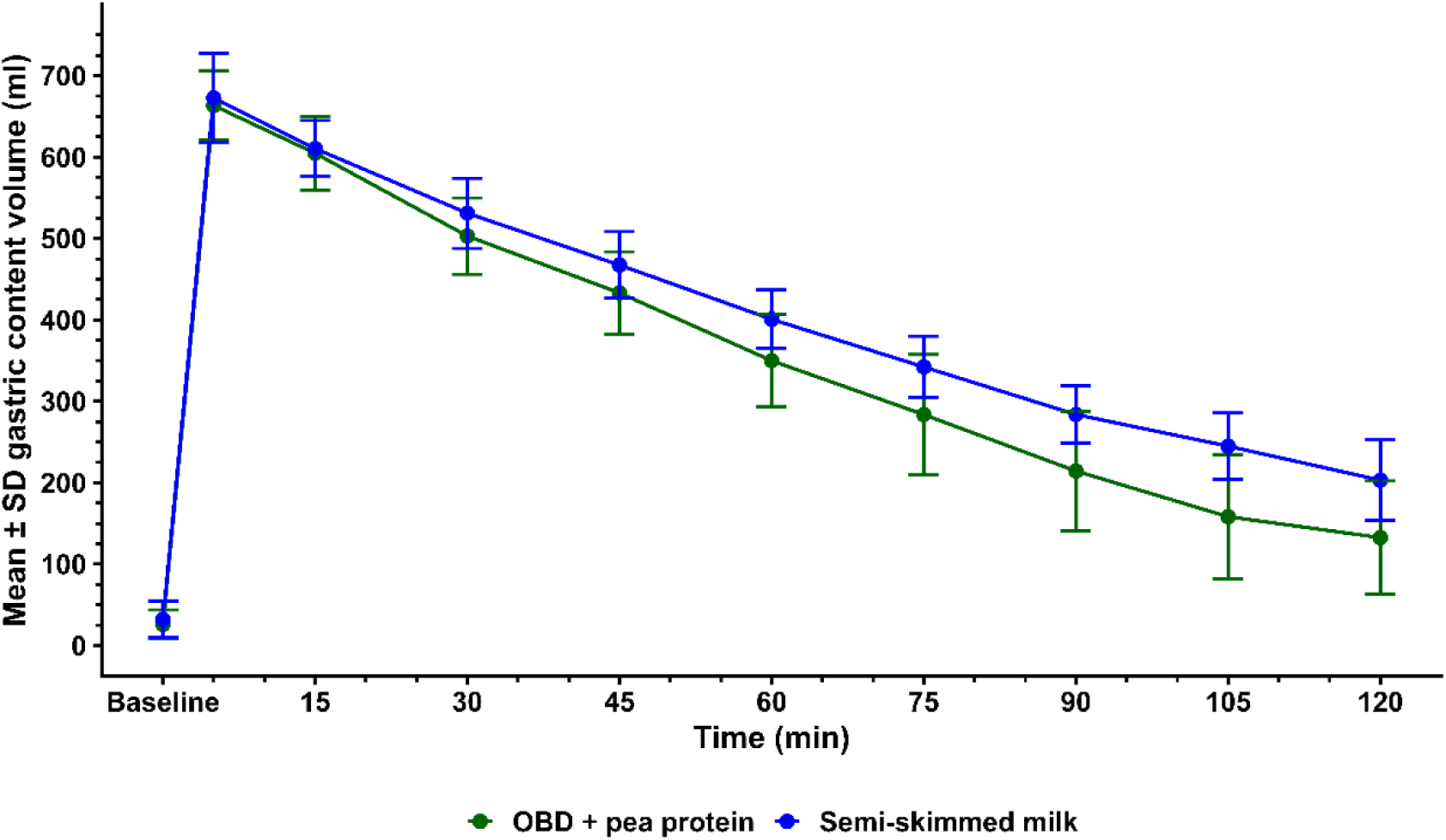
Mean±SD gastric content volume over time for OBD+Pea and Milk (n=12). Baseline represents the empty stomach after an overnight fast. T=0 min is the start of consumption. Postprandial volumes were analyzed using linear mixed models with the treatment*time interaction as a fixed factor and baseline volume as a covariate.

Coagulation: Visual inspection of all gastric MRI time series showed little signs of protein coagulation, as illustrated in Supplementary figure 2. In line with this, the MRI image grey value thresholding analysis did not show any differences in the number of voxels classified as Liquid (higher grey value) or more Solid (lower grey value) between Milk and OBD+Pea at any time point (**Supplementary figure 6**). Image texture (**Supplementary figure 7**): All texture metrics changed over time (all P<0.001). Joint variance was higher for Milk from 5 till 30 min (all P<0.02). It increased over time for OBD+Pea, while for Milk it remained at the same level until 60 min and then started to increase along with OBD+Pea. Contrast increased over time for both drinks but was higher for Milk from 5 until 45 min (all P<0.02) and at 75 min (P=0.027). Coarseness increased over time but faster for OBD+Pea from 60 min onwards. As a results, it was higher for OBD+Pea from 90 min onwards (all P<0.004). Busyness decreased over time and was consistently higher for Milk (all P<0.004).

### FGF21

FGF21 curves are shown in **Supplementary figure 8**. There was a significant effect of treatment (P<0.001). Averaged over time, FGF21 concentrations were highest following OBD (estimated marginal mean 91, 95% CI (85, 97), intermediate following OBD+Pea (78, 95% CI (72, 84), and lowest following Milk (70, 95% CI (64, 76). Compared with Milk, OBD elicited higher FGF21 concentrations (mean difference: 21, 95% CI (15, 28), P <0.001), whereas OBD+Pea resulted in a more modest increase (mean difference: 8, 95% CI (2, 15), P=0.023). FGF21 concentrations were also higher following OBD compared with OBD+Pea (mean difference: 13, 95% CI (7, 19), P<0.001).

The overall treatment by time interaction was not significant (P=0.44). However, compared with Milk, FGF21 concentrations were higher following OBD at 120 min (mean difference: 31, 95% CI (9, 53), P = 0.018), 150 min (43, 95% CI (21, 66), P < 0.001), 180 min (41, 95% CI (19, 63), P=0.001), and 210 min (35, 95% CI (13, 57), P=0.006). No other pairwise comparisons reached statistical significance (all P ≥ 0.07).

### Subjective ratings

Plots of the subjective ratings over time are shown in **Supplementary Figure 9**. Nausea, stomach complaints, fullness and thirst, did not show treatment differences or treatment by time interaction. Nausea and stomach complaint scores were very low for all treatments (mean score per timepoint ranged between 0 and 4, for both). There was an overall effect of treatment for hunger with slightly higher postprandial hunger for OBD compared to OBD+Pea (mean difference: 8.2, 95% CI (4.6, 11.7), P<0.001) and Milk (mean difference: 5.1, 95% CI (1.5, 8.7), P=0.015).

## Discussion

This study examined plasma amino acid concentrations, gastric behavior, insulin and glucose concentrations, FGF21 concentrations, and subjective ratings related to appetite and wellbeing after consumption of milk, an oat-based drink and an oat-based drink with added pea protein. Milk consumption resulted in the highest postprandial total and essential amino acid concentrations, OBD+Pea induced intermediate responses, and OBD the lowest. Both OBD+Pea and Milk emptied from the stomach in an approximately linear manner. However, Milk emptied significantly slower than OBD+Pea after the first 60 min. Insulin and glucose excursions were higher for the two OBDs compared to Milk during the first 15-60 min. FGF21 concentrations were higher for OBD compared to Milk from 120 to 210 min. Hunger ratings were slightly higher for OBD compared to OBD+Pea and Milk.

### Amino acid responses

The three drinks differed in nutritional composition (Supplementary Table 1), as they are commercially available products rather than matched research products formulated for specific comparisons. Notably, they varied in protein content, with milk containing the highest amount, followed by OBD+Pea and OBD. This was reflected in the total amino acid curves, with higher protein content leading to higher total amino acid concentrations. Furthermore, differences in protein quality likely also contributed to the observed differences although products should be tested at equal protein content to be able to draw conclusions on the effect of protein quality alone, which was not within the scope of the current study.

In addition, the EAA curves mirrored both the differences in protein quantity and in protein quality (amino acid composition) between the test products. Milk, like most animal products, contains higher concentrations of EAA compared to plant-based products, with milk protein having a more balanced EAA profile compared to plant-based protein [44]. Oat protein present in the oat-based drinks, on the other hand, is limited in lysine [45]. The combination with pea protein in the OBD+Pea improves its amino acid profile contributing to the higher EAA concentrations, since pea has a complementary amino acid composition, with higher relative lysine content, while oat protein compensates the lack of sulphur-containing EAAs (methionine and cysteine) of pea protein [46].

### Gastric behavior

After the first hour, Milk emptied slower than OBD+Pea. This can be explained by the higher caloric density and protein content of the milk; it is well established that higher caloric density leads to slower gastric emptying [47–49]. Part of this may be caused by the difference in protein content between the two drinks (Milk 28 g / OBD+Pea 13 g). For iso-caloric loads protein is generally the most satiating macronutrient (e.g. [50–52]. This is thought to be mediated by stronger appetite-related gut hormone responses [53–55], which in turn affect gastric emptying rate. Accordingly, higher protein content has been associated with slower gastric emptying and greater satiety-related gut hormone responses [56,57]. The different amino acid composition might also have influenced gastric emptying and related processes, but to date there is limited evidence for this [58]. Finally, we note that the observed difference in gastric emptying may not be real-life relevant, since hunger ratings were only slightly higher for OBD compared to OBD+Pea and milk, and no significant differences were observed between OBD+Pea and milk, suggesting that the faster gastric emptying of OBD+Pea was hardly noticed by participants in terms of more hunger or less experienced fullness. This aligns with the finding that, unlike gastric emptying rate, appetite sensations depend more on drink viscosity (which was similar here) than on caloric density [49]. Moreover, experienced appetite differences after liquid meals may not translate into differences in subsequent food intake [49,59].

There was no apparent protein coagulation for either drink, as reflected in the absence of clear structure formation, assessed both by visual inspection and thresholding analysis of the gastric MRI images. This is putatively due to the UHT treatment of the milk which has been shown to lead to weaker curds [60–62]. In addition, a larger milk load such as used here (600 ml) requires more gastric acid to reach the low pH needed for pepsin action. E.g. for a 250-ml UHT milk load clear coagulation has been observed with MRI from ∼30 min onwards [10]. For OBD+Pea no coagulation was expected. This is supported by the lack of pea-protein coagulation in an MRI study using 420-ml drinks containing 20 g pea protein isolate [14]. Still, the image texture analysis showed increasing Contrast over time for both drinks with higher values for Milk from 5 until 45 minutes. Coarseness also increased over time for both drinks but was higher for OBD+Pea from 90 min onwards. Such increases have also been reported for UHT milk [10], pasteurized skimmed milk [43] and pea-protein drinks [14]. Joint variance was higher for Milk from 5 – 30 minutes and then increased over time along with OBD+Pea; on this metric the two drinks converged. A decline in Busyness was also seen for pea-protein drinks [14] and pasteurized skim milk [43], but there was no difference between the two drinks in this study. The differences in Contrast and Coarseness suggest that there were relatively subtle structural differences between the digesta of the two drinks over time related to small-scale protein-related aggregation, but their exact nature would require *in vitro* validation.

### Glycemic responses

Compared to Milk the two OBDs induced higher insulin and glucose concentrations. Milk has the highest sugar content but the OBDs had the highest carbohydrate content (Supplementary table 1). The lower glucose and insulin response for Milk can be attributed to the low glycemic index of lactose [22], which is the main sugar present in bovine milk. At the same time, although the regular OBD used in this study is ‘sugar-free’ its carbohydrate fraction was high glycemic compared to Milk. This aligns with the observation that plant-based drinks marketed as milk substitutes have a higher glycemic index than milk [22,23]. Our findings corroborate that the oat starches in oat-based drinks and their predominant breakdown products dextrins and maltodextrins have a high digestibility, in contrast to less-processed oat products [63], which leads to their higher glycemic index. The generally slower GE of Milk compared to the OBDs could further contribute to the lower postprandial glucose concentrations although it is likely that the differences in carbohydrate source are the main driver of the observed lower glycemic response. Note that the glucose peak for milk was lower compared to that for the OBDs, even though glucose peaked at the same time (15 min) for all three drinks. Insulin concentrations reached a peak at 15 min for Milk, while the peak occurred at 30 min for the OBDs. This could indicate that milk protein might have started to stimulate insulin secretion before increasing glucose concentrations did. Although glucose is the primary driver of insulin secretion, dietary protein is also known to exert insulinotropic effects. In particular, branched-chain amino acids, especially leucine, as well as other amino acids such as arginine and alanine, can stimulate insulin release and thereby contribute to postprandial insulin responses [21,64]. On the other hand, the insulin peak for the OBDs at 30 min, after the glucose peak at 15 min, suggests that this peak constitutes a response to the rise in glucose. The results for Milk also showed that the lowest glucose concentration was lower than for the two OBDs, despite a lower insulin response. These findings indicate that milk might be a better option in individuals in which glycemic regulation is compromised e.g. because of pre-diabetics or type 2 diabetes, but further studies are needed to confirm this.

### FGF21

Higher circulating FGF21 concentrations are associated with lower protein intake [24,28]. This could explain why FGF21 concentrations differed significantly between all drinks, with higher concentrations corresponding to lower protein content of the intervention. Methionine restriction specifically has been shown to lead to higher FGF21 concentrations [28,65] which could further explain why FGF21 was lower after milk ingestion compared to the OBDs, since animal protein is generally richer in methionine compared to plant protein. To further discriminate between the effect of protein quantity and protein quality further studies using equal protein loads should be performed. In addition, the results show high standard deviations, suggesting large interpersonal differences.

## Conclusion

In conclusion, the enrichment of an oat-based drink with pea protein partially improved amino acid composition but did not fully replicate the one of bovine milk and was associated with faster gastric emptying. In addition, both OBDs induced a higher early postprandial glucose and insulin response compared to bovine milk, because of their greater and high-glycemic carbohydrate content. These findings highlight that protein enrichment of plant-based milk alternatives can improve amino acid composition, but that there are still relevant differences in digestive and metabolic responses between bovine milk and plant-based alternatives.

## Data Availability

All data presented in the manuscript are available upon reasonable request to the authors.

## Acknowledgements

We thank Meike Snoeren for her help with data collection and Nhien Ly for performing the FGF21 laboratory analysis. The use of the 3T MRI was made possible by WUR Shared Research Facilities.

## Author contributions are as follows

PAMS and TL designed the research; LL and SC conducted the research; GdG and MB performed the amino acid analyses, PS and SC analyzed data; TL, PS,SC and LL interpreted the results; SC, LL and PS wrote the paper. PS had primary responsibility for final content. All authors provided feedback on the draft manuscript and read and approved the final manuscript.

## Data Availability

Data described in the manuscript and analytic code will be made available for research use upon reasonable request to the corresponding author.

## Declaration of Generative AI and AI-assisted technologies in the writing process

During the writing process the authors occasionally used Generative AI and AI-assisted technologies (enterprise version of Microsoft Copilot) to improve phrasing and conciseness. The authors take full responsibility for the final text.

## Abbreviations

Please define all nonstandard abbreviations and group designations (e.g. CON, HP) used at first mention in the abstract and text. Alphabetically list and define them.

AA: amino acid
BCAA: branched-chain amino acid
EAA: essential amino acids
EDTA: ethylenediaminetetraacetic acid
FGF21: fibroblast Growth Factor 21
HILIC: hydrophilic interaction liquid chromatography
iAUC: incremental area under the curve
MRI: magnetic resonance imaging
UHT: ultra-high temperature
UPLC-MS/MS: ultrahigh pressure liquid chromatography coupled to tandem mass spectrometry

