## Supplementary Materials for "Postprandial amino acid profiles and gastric behavior after consumption of milk and plant-based alternatives: a randomized cross-over study in healthy males"

### Contents


- Participant flow chart
- Drink composition
- Gastric content: MRI illustration
- Amino acid analysis details
- Total AA and EAA tables
- Individual amino acid plots
- Glucose and Insulin tables
- Gastric content: individual volume plots
- Gastric content: image texture metric plots
- FGF21 plot
- Subjective rating plots

Participant flow chart

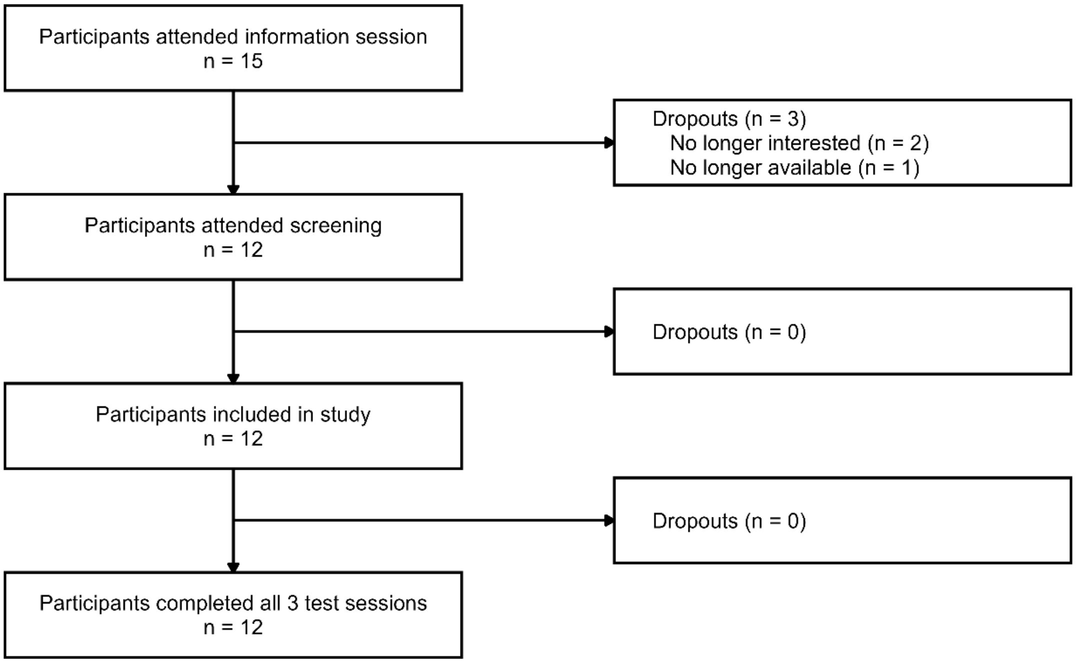


*Supplementary Figure 1. Participant flow chart.*

Drink composition

**Supplementary table 1.** Drink composition per 100 ml and per 750-ml serving.

| | **Oat-based drink** | | **Oat-based drink with added pea protein** | | **Semi-skimmed milk** | |
|---|---|---|---|---|---|---|
| | *100 ml* | *750 ml* | *100 ml* | *750 ml* | *100 ml* | *750 ml* |
| **Energy (kJ/kcal)** | 183/44 | 1373/330 | 194/46 | 1455/345 | 204/48 | 1530/360 |
| **Fat (g)** | 1.8 | 13.5 | 1.5 | 11.3 | 1.5 | 11.3 |
| **Of which saturated (g)** | 0.2 | 1.5 | 0.2 | 1.5 | 1.1 | 8.3 |
| **Carbohydrates (g)** | 5.7 | 42.8 | 6.2 | 46.5 | 4.8 | 36 |
| **Of which sugars (g)** | 0 | 0 | 3.5 | 26.3 | 4.8 | 36 |
| **Protein (g)** | 0.7 | 5.25 | 1.7 | 12.8 | 3.7 | 27.8 |
| **Salt (g)** | 0.12 | 0.9 | 0.18 | 1.35 | 0.13 | 0.98 |
| **Calcium (mg)** | 120 | 900 | 120 | 900 | 120 | 900 |

Amino acid analysis details

*Abbreviations:*

| | |
|---|---|
| ESI | Electrospray ionization |
| UPLC | ultrahigh pressure liquid chromatography |
| HILIC | Hydrophilic interaction liquid chromatography |
| MS/MS | triple quad mass spectrometer |
| MRM | multiple reaction monitoring |

*Chemicals:*

The following chemicals were purchased: Water ULC/MS (Biosolve, 00232141B1BS), Acetonitril ULC/MS (Biosolve, 0001204101BS), Formic acid ULC/MS (Biosolve, 0006914143BS), Ammonium formate (), Methanol ULC/MS (Biosolve, 0013684101BS), Isopropanol ULC/MS (Biosolve, 0016264102BS), L-Alanine (Sigma, A7627), L-Arginine Hydrochloride (Sigma, A5131), L-Asparagine (Sigma, A0884), L-Aspartic Acid (Sigma, A9256), L-Cysteine hydrochloride (Sigma, C1276), L-Cystine (Sigma, C8755), L-Glutamic Acid (Sigma, G1251), L-Glutamine (Sigma, G3126), Glycine (Sigma, G7126), L-Histidine hydrochloride (Sigma, H8125), trans-4-Hydroxy-L-proline (Sigma, H5534), L-Isoleucine (Sigma, I2752), L-Leucine (Sigma, L8000), L-Lysine hydrochloride (Sigma, L5626), L-Methionine (Sigma, M9625), L-Phenylalanine (Sigma, P2126), L-Proline (Sigma, P0380), L-Serine (Sigma, S4500), L-Threonine (Sigma, T8625), L-Tryptophan (Sigma, T0254), L-Tyrosine (Sigma, T3754), L-Valine (Sigma, V0500), Taurine (Sigma, T0625), Citrulline (Sigma, C7629), Alpha-aminobutyric acid (AABA) (Sigma, 162663), Ornithine (Sigma, O2375) and Norvaline (Sigma, 53721).

For every compound an individual 50 mM stock solution was prepared (except for L-glutamine and L-alanine 100 mM) in water. The calibrator with the highest concentration was prepared by pipetting 100 µl of every stock solution in a 10 ml volumetric flask and filling this up with water. 7 other calibrators were prepared freshly before analysis by diluting calibrators 1:1 with water.

The internal standard mixture containing $^{13}C_3$-$^{15}N$-L-Alanine, $^{13}C_6$-$^{15}N_4$-L-Arginine, $^{13}C_4$-$^{15}N_2$-L-Asparagine, $^{13}C_4$-$^{15}N$-L-Aspartic acid, $^{13}C_6$-$^{15}N_2$-L-Cystine, $^{13}C_5$-$^{15}N$-L-Glutamic acid, $^{13}C_5$-$^{15}N_2$-L-Glutamine, $^{13}C_2$-$^{15}N$-Glycine, $^{13}C_6$-$^{15}N_3$-L-Histidine, $^{13}C_6$-$^{15}N$-L-Isoleucine, $^{13}C_6$-$^{15}N$-L-Leucine, $^{13}C_6$-$^{15}N_2$-L-Lysine, $^{13}C_5$-$^{15}N$-L-Methionine, $^{13}C_9$-$^{15}N$-L-Phenylalanine, $^{13}C_5$-$^{15}N$-L-Proline, $^{13}C_3$-$^{15}N$-L-Serine, $^{13}C_4$-$^{15}N$-L-Threonine, $^{13}C_{11}$-$^{15}N_2$-L-Tryptophan, $^{13}C_9$-$^{15}N$-L-Tyrosine and $^{13}C_5$-$^{15}N$-L-Valine was purchased via Cambridge Isotope Laboratories, Inc. (MSK-CAA-1). According to the provided protocol, adding 1 ml of water resulted the concentration to be 2.5 mM (except for $^{13}C_6$-$^{15}N_2$-L-Cystine 1.25 mM). This solution was diluted 50 times with water to obtain the internal standard working solution.

*Sample preparation:*

40 µl internal standard working solution was added to 40 µl of standard or QC sample or plasma in a 1.5 ml Eppendorf tube. 280 µl of mobile phase A was added (see below for its composition) and the mixture was vortexed. After centrifuging the mixture for 15 min at 13.000 rpm, the supernatant was transferred into a 96-well plate and was ready for analysis.

*Method:*

The quantitative analysis of amino acids was performed by ultrahigh pressure liquid chromatography (UPLC) coupled to triple quad mass spectrometer (MS/MS). Based on a hydrophilic interaction liquid chromatography (HILIC) method published by Prinsen et al. (J Inherit Metab Dis. 2016 Sep;39(5):651-660. doi: 10.1007/s10545-016-9935-z. PMID: 27099181).

Chromatographic separation was obtained on an ACQUITY UPLC BEH Amide Column, 130Å, 1.7 µm, 2.1 mm X 100 mm (Waters) using a ACQUITY UPLC BEH Amide VanGuard Pre-column, 130Å, 1.7 µm, 2.1 mm X 5 mm (Waters). The autosampler temperature was set at 5°C, the column temperature was maintained at 35 °C and the injection volume was 2 µl. Mobile phase A consists of 10 mM ammonium formate in 85% ACN containing 0.15% formic acid and Mobile phase B consist of 10 mM ammonium formate in water containing 0.15% formic acid. The following linear gradient was used at a flow of 0.4 ml/min. 100% A was maintained for the first 6 minutes. From 6.0 to 6.1 minutes, the percentage A was reduced to 94.1%. From 6.1 to 10.0 minutes, the percentage A was reduced to 82.4%. the percentage A was further reduced till 70.6% at 12.0 minutes. And from minute 12.0 till the end of the runtime (18 minutes) the percentage A was increased to 100%. With the correction of an internal standard for every compound, linear calibration curves were fitted with a 1/x^2 weighting factor. Coefficients of variation of inter batch QC samples are below 15% for amino acids with their own similar internal standard (except for aspartic acid, glutamic acid and lysine).

*Instrument:*

An Acquity H-class Plus UPLC (Waters Chromatography Europe BV, Etten-Leur, the Netherlands) coupled to a Xevo TQ-S micro tandem mass spectrometer (Waters Chromatography Europe BV, Etten-Leur, the Netherlands) was used. Electrospray ionization (ESI) was operated in the positive mode and using multiple reaction monitoring (MRM), the amino acids were analyzed. An overview of the compounds with their retention time, parent and precursor m/z, cone voltage and collision energy is shown in the **Table** below. Data was processed using MassLynx v4.2 software.

**Table.** Overview parameters per compound.

| | **Compound** | **Retention time (min)** | **Parent m/z** | **Precursor m/z** | **Cone voltage (V)** | **Collision energy (V)** | **Calibration range (µM)** | **Internal standard used** |
|---|---|---|---|---|---|---|---|---|
| 1 | Alanine | 6.5 | 90.2 | 44.2 | 20 | 9 | 8 - 1000 | $^{13}C_3$-$^{15}N$-L-Alanine |
| 2 | Alpha-aminobutyric acid | 4.9 | 104.3 | 58.3 | 20 | 12 | 4 - 500 | $^{13}C_5$-$^{15}N$-L-Methionine |
| 3 | Arginine | 11.2 | 175.2 | 70.2 | 30 | 30 | 4 - 500 | $^{13}C_6$-$^{15}N_4$-L-Arginine·HCl |
| 4 | Asparagine | 9.2 | 133.2 | 74.2 | 10 | 20 | 4 - 500 | $^{13}C_4$-$^{15}N_2$-L-Asparagine |
| 5 | Aspartic acid | 10.7 | 134.08 | 74.21 | 20 | 18 | 4 - 500 | $^{13}C_4$-$^{15}N$-L-Aspartic acid |
| 6 | Citrulline | 9.6 | 176.3 | 159.9 | 20 | 10 | 4 - 500 | $^{13}C_6$-$^{15}N_4$-L-Arginine·HCl |
| 7 | Cysteine | 4.9 | 122.3 | 76.1 | 15 | 10 | 4 - 500 | $^{13}C_6$-$^{15}N_2$-L-Cystine |
| 8 | Cystine | 12.6 | 241.3 | 74.1 | 10 | 35 | 4 - 500 | $^{13}C_6$-$^{15}N_2$-L-Cystine |

| | Compound | Retention time (min) | Parent m/z | Precursor m/z | Cone voltage (V) | Collision energy (V) | Calibration range (µM) | Internal standard used |
|---|---|---|---|---|---|---|---|---|
| 9 | Glutamic acid | 9.6 | 148.09 | 84.2 | 16 | 20 | 4 - 500 | $^{13}C_5$-$^{15}N$-L-Glutamic acid |
| 10 | Glutamine | 8.9 | 147.0 | 84.2 | 18 | 18 | 8 - 1000 | $^{13}C_5$-$^{15}N_2$-L-Glutamine |
| 11 | Glycine | 8.0 | 76.2 | 30.2 | 15 | 8 | 4 - 500 | $^{13}C_2$-$^{15}N$-Glycine |
| 12 | Histidine | 11.3 | 156.2 | 110.4 | 18 | 20 | 4 - 500 | $^{13}C_6$-$^{15}N_3$-L-Histidine·HCl |
| 13 | Hydroxyproline | 6.6 | 132.11 | 86.3 | 44 | 14 | 4 - 500 | $^{13}C_5$-$^{15}N$-L-Proline |
| 14 | Isoleucine | 2.8 | 132.3 | 86.3 | 18 | 18 | 4 - 500 | $^{13}C_6$-$^{15}N$-L-Isoleucine |
| 15 | Leucine | 2.5 | 132.3 | 86.3 | 18 | 18 | 4 - 500 | $^{13}C_6$-$^{15}N$-L-Leucine |
| 16 | Lysine | 11.5 | 147.5 | 84.5 | 18 | 20 | 4 - 500 | $^{13}C_6$-$^{15}N_2$-L-Lysine·2HCl |
| 17 | Methionine | 3.2 | 150.4 | 104.4 | 18 | 12 | 4 - 500 | $^{13}C_5$-$^{15}N$-L-Methionine |
| 18 | Norvaline | 3.2 | 118.3 | 72.4 | 10 | 12 | 4 - 500 | $^{13}C_5$-$^{15}N$-L-Valine |
| 19 | Ornithine | 11.6 | 133.5 | 70.4 | 20 | 18 | 4 - 500 | $^{13}C_6$-$^{15}N_4$-L-Arginine·HCl |
| 20 | Phenylalanine | 2.4 | 166.4 | 120.4 | 20 | 15 | 4 - 500 | $^{13}C_9$-$^{15}N$-L-Phenylalanine |
| 21 | Proline | 4.0 | 116.4 | 70.3 | 15 | 15 | 4 - 500 | $^{13}C_5$-$^{15}N$-L-Proline |
| 22 | Serine | 9.0 | 106.3 | 60.3 | 20 | 15 | 4 - 500 | $^{13}C_3$-$^{15}N$-L-Serine |
| 23 | Taurine | 4.1 | 126.2 | 44.4 | 15 | 19 | 4 - 500 | $^{13}C_5$-$^{15}N$-L-Proline |
| 24 | Threonine | 7.6 | 120.3 | 74.3 | 15 | 12 | 4 - 500 | $^{13}C_4$-$^{15}N$-L-Threonine |
| 25 | Tryptophan | 2.4 | 205.2 | 188.2 | 20 | 15 | 4 - 500 | $^{13}C_{11}$-$^{15}N_2$-L-Tryptophan |
| 26 | Tyrosine | 4.0 | 182.4 | 136.4 | 18 | 18 | 4 - 500 | $^{13}C_9$-$^{15}N$-L-Tyrosine |
| 27 | Valine | 3.8 | 118.2 | 72.2 | 18 | 12 | 4 - 500 | $^{13}C_5$-$^{15}N$-L-Valine |
| 28 | $^{13}C_3$-$^{15}N$-L-Alanine | 6.5 | 94.20 | 47.2 | 20 | 9 | | |
| 29 | $^{13}C_6$-$^{15}N_4$-L-Arginine | 11.2 | 185.2 | 75.2 | 30 | 30 | | |
| 30 | $^{13}C_4$-$^{15}N_2$-L-Asparagine | 9.2 | 139.2 | 77.2 | 10 | 20 | | |
| 31 | $^{13}C_4$-$^{15}N$-L-Aspartic acid | 10.7 | 139.08 | 77.21 | 20 | 18 | | |
| 32 | $^{13}C_6$-$^{15}N_2$-L-Cystine | 12.6 | 249.3 | 77.1 | 10 | 35 | | |
| 33 | $^{13}C_5$-$^{15}N$-L-Glutamic acid | 9.6 | 154.09 | 89.2 | 16 | 20 | | |
| 34 | $^{13}C_5$-$^{15}N_2$-L-Glutamine | 8.9 | 154.00 | 89.20 | 18 | 18 | | |
| 35 | $^{13}C_2$-$^{15}N$-Glycine | 8.1 | 79.2 | 32.2 | 15 | 8 | | |
| 36 | $^{13}C_6$-$^{15}N_3$-L-Histidine | 11.3 | 165.2 | 118.4 | 18 | 20 | | |
| 37 | $^{13}C_6$-$^{15}N$-L-Leucine | 2.5 | 139.3 | 92.3 | 18 | 18 | | |
| 38 | $^{13}C_6$-$^{15}N$-L-Isoleucine | 2.8 | 139.3 | 92.3 | 18 | 18 | | |
| 39 | $^{13}C_6$-$^{15}N_2$-L-Lysine | 11.5 | 155.5 | 90.5 | 18 | 20 | | |
| 40 | $^{13}C_5$-$^{15}N$-L-Methionine | 3.2 | 156.4 | 109.4 | 18 | 12 | | |
| 41 | $^{13}C_9$-$^{15}N$-L-Phenylalanine | 2.4 | 176.4 | 129.4 | 20 | 15 | | |
| 42 | $^{13}C_5$-$^{15}N$-L-Proline | 4.0 | 122.4 | 75.3 | 15 | 15 | | |
| 43 | $^{13}C_3$-$^{15}N$-L-Serine | 9.0 | 110.3 | 63.3 | 20 | 15 | | |
| 44 | $^{13}C_4$-$^{15}N$-L-Threonine | 7.6 | 125.3 | 78.3 | 15 | 12 | | |
| 45 | $^{13}C_{11}$-$^{15}N_2$-L-Tryptophan | 2.4 | 218.2 | 200.2 | 20 | 15 | | |
| 46 | $^{13}C_9$-$^{15}N$-L-Tyrosine | 4.0 | 192.4 | 145.4 | 18 | 18 | | |
| 47 | $^{13}C_5$-$^{15}N$-L-Valine | 3.8 | 124.2 | 77.2 | 18 | 12 | | |

Gastric content: MRI illustration

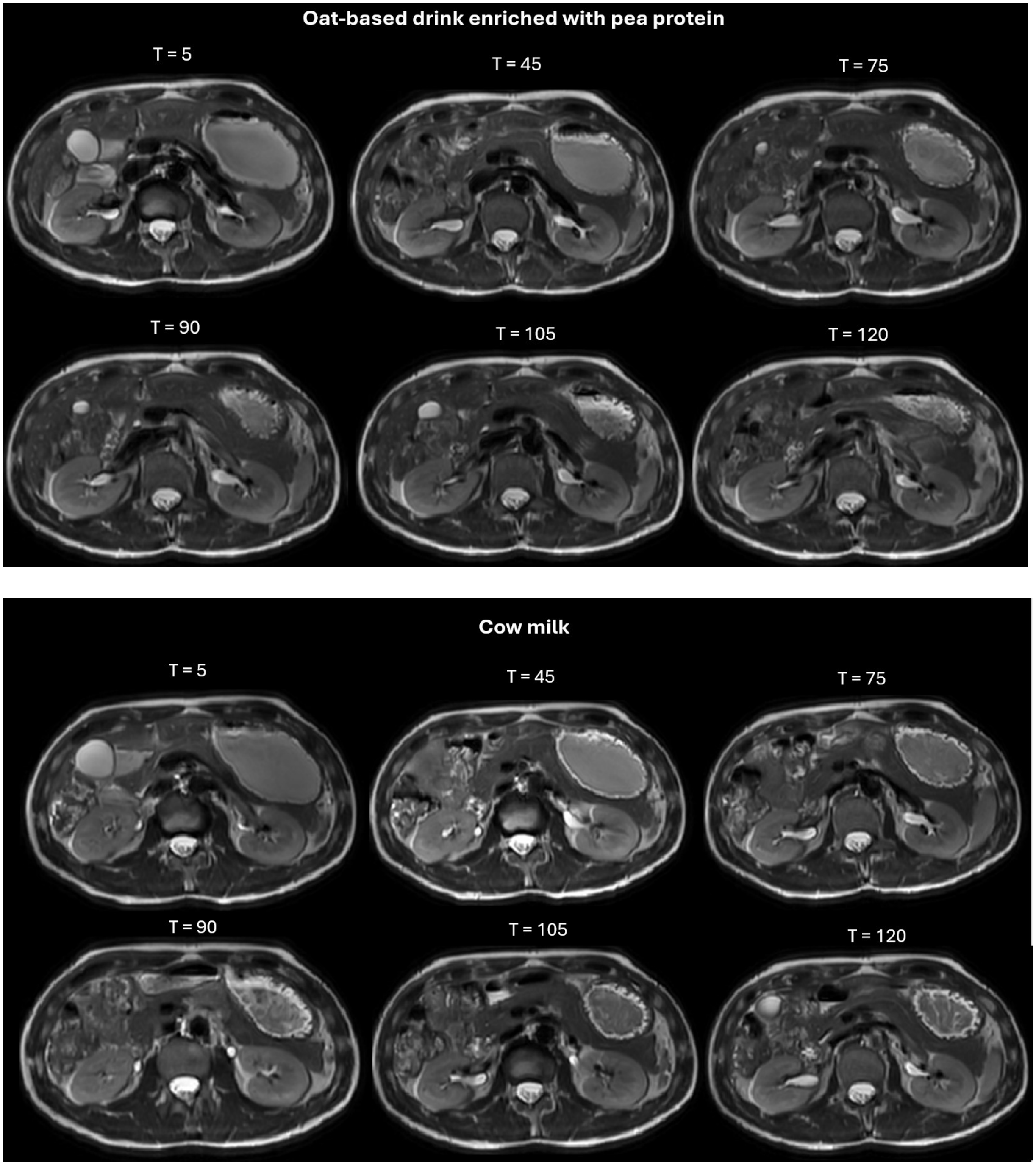


*Supplementary Figure 2. Illustration of stomach scan time series for the two treatments with MRI measurements (one participant). Shown are axial cross-sections through the abdomen.*

Total AA and EAA results tables

**Supplementary Table 2**. Treatment differences in estimated marginal mean total AA concentration (95% CI) in µM/l per timepoint, adjusted for baseline concentration.

| Timepoint (min) | Contrast | Mean difference in total AA concentration (95% CI) | P-value (tukey-corrected) |
|---|---|---|---|
| T=15 | OBD - (OBD+Pea) | -333 (-617, -50) | **0.016** |
| | OBD - Milk | -844 (-1128, -561) | **<0.001** |
| | (OBD+Pea) - Milk | -511 (-794, -228) | **<0.001** |
| T=30 | OBD - (OBD+Pea) | -369 (-652, -86) | **0.007** |
| | OBD - Milk | -1052 (-1335, -768) | **<0.001** |
| | (OBD+Pea) - Milk | -683 (-966, -400) | **<0.001** |
| T=45 | OBD - (OBD+Pea) | -602 (-885, -319) | **<0.001** |
| | OBD - Milk | -1183 (-1466, -899) | **<0.001** |
| | (OBD+Pea) - Milk | -580 (-863, -297) | **<0.001** |
| T=60 | OBD - (OBD+Pea) | -490 (-773, -207) | **<0.001** |
| | OBD - Milk | -1027 (-1311, -744) | **<0.001** |
| | (OBD+Pea) - Milk | -538 (-821, -255) | **<0.001** |
| T=75 | OBD - (OBD+Pea) | -535 (-825, -246) | **<0.001** |
| | OBD - Milk | -1330 (-1613, -1047) | **<0.001** |
| | (OBD+Pea) - Milk | -795 (-1084, -505) | **<0.001** |
| T=90 | OBD - (OBD+Pea) | -392 (-675, -109) | **0.003** |
| | OBD - Milk | -1169 (-1452, -885) | **<0.001** |
| | (OBD+Pea) - Milk | -777 (-1060, -494) | **<0.001** |
| T=105 | OBD - (OBD+Pea) | -509 (-792, -226) | **<0.001** |
| | OBD - Milk | -920 (-1204, -637) | **<0.001** |
| | (OBD+Pea) - Milk | -411 (-694, -128) | **0.002** |
| T=120 | OBD - (OBD+Pea) | -192 (-475, 91) | 0.248 |
| | OBD - Milk | -888 (-1178, -598) | **<0.001** |
| | (OBD+Pea) - Milk | -696 (-985, -406) | **<0.001** |
| T=150 | OBD - (OBD+Pea) | -165 (-448, 118) | 0.356 |
| | OBD - Milk | -904 (-1187, -620) | **<0.001** |
| | (OBD+Pea) - Milk | -739 (-1022, -456) | **<0.001** |
| T=180 | OBD - (OBD+Pea) | -180 (-463, 103) | 0.293 |
| | OBD - Milk | -839 (-1122, -555) | **<0.001** |
| | (OBD+Pea) - Milk | -658 (-941, -375) | **<0.001** |
| T=210 | OBD - (OBD+Pea) | -82 (-365, 201) | 0.773 |
| | OBD - Milk | -684 (-968, -401) | **<0.001** |
| | (OBD+Pea) - Milk | -602 (-885, -319) | **<0.001** |
| T=240 | OBD - (OBD+Pea) | -141 (-424, 142) | 0.468 |
| | OBD - Milk | -414 (-697, -130) | **0.002** |
| | (OBD+Pea) - Milk | -272 (-555, 11) | **0.062** |
| T=300 | OBD - (OBD+Pea) | 18 (-265, 301) | 0.988 |

| | | |
|---|---|---|
| OBD - Milk | -218 (-502, 65) | 0.167 |
| (OBD+Pea) - Milk | -237 (-520, 46) | 0.122 |

**Supplementary Table 3**. Treatment differences in estimated marginal mean essential AA concentration (95% CI) in µM/l per timepoint, adjusted for baseline concentration.

| **Timepoint (min)** | **Contrast** | **Mean difference in EAA concentration (95% CI)** | **P-value (tukey-corrected)** |
|---|---|---|---|
| T=15 | OBD - (OBD+Pea) | -163 (-284, -41) | **0.005** |
| | OBD - Milk | -435 (-556, -313) | **<0.001** |
| | (OBD+Pea) - Milk | -272 (-394, -151) | **<0.001** |
| T=30 | OBD - (OBD+Pea) | -178 (-300, -57) | **0.002** |
| | OBD - Milk | -530 (-652, -409) | **<0.001** |
| | (OBD+Pea) - Milk | -352 (-474, -231) | **<0.001** |
| T=45 | OBD - (OBD+Pea) | -260 (-382, -139) | **<0.001** |
| | OBD - Milk | -541 (-662, -419) | **<0.001** |
| | (OBD+Pea) - Milk | -281 (-402, -159) | **<0.001** |
| T=60 | OBD - (OBD+Pea) | -225 (-346, -103) | **<0.001** |
| | OBD - Milk | -501 (-623, -380) | **<0.001** |
| | (OBD+Pea) - Milk | -276 (-398, -155) | **<0.001** |
| T=75 | OBD - (OBD+Pea) | -233 (-357, -108) | **<0.001** |
| | OBD - Milk | -680 (-801, -558) | **<0.001** |
| | (OBD+Pea) - Milk | -447 (-572, -323) | **<0.001** |
| T=90 | OBD - (OBD+Pea) | -179 (-301, -58) | **0.002** |
| | OBD - Milk | -550 (-672, -429) | **<0.001** |
| | (OBD+Pea) - Milk | -371 (-492, -250) | **<0.001** |
| T=105 | OBD - (OBD+Pea) | -205 (-327, -84) | **<0.001** |
| | OBD - Milk | -504 (-626, -383) | **<0.001** |
| | (OBD+Pea) - Milk | -299 (-421, -178) | **<0.001** |
| T=120 | OBD - (OBD+Pea) | -102 (-223, 20) | 0.120 |
| | OBD - Milk | -470 (-595, -346) | **<0.001** |
| | (OBD+Pea) - Milk | -368 (-493, -244) | **<0.001** |
| T=150 | OBD - (OBD+Pea) | -88 (-210, 33) | 0.203 |
| | OBD - Milk | -480 (-601, -358) | **<0.001** |
| | (OBD+Pea) - Milk | -392 (-513, -270) | **<0.001** |
| T=180 | OBD - (OBD+Pea) | -114 (-236, 7) | 0.070 |
| | OBD - Milk | -443 (-565, -322) | <0.001 |
| | (OBD+Pea) - Milk | -329 (-450, -207) | <0.001 |
| T=210 | OBD - (OBD+Pea) | -36 (-158, 85) | 0.762 |
| | OBD - Milk | -382 (-503, -260) | **<0.001** |
| | (OBD+Pea) - Milk | -345 (-467, -224) | **<0.001** |
| T=240 | OBD - (OBD+Pea) | -45 (-166, 77) | 0.660 |
| | OBD - Milk | -258 (-380, -137) | **<0.001** |
| | (OBD+Pea) - Milk | -213 (-335, -92) | **<0.001** |
| T=300 | OBD - (OBD+Pea) | -24 (-146, 97) | 0.887 |
| | OBD - Milk | -175 (-296, -53) | 0.002 |
| | (OBD+Pea) - Milk | -151 (-272, -29) | 0.010 |

### Amino acid plots

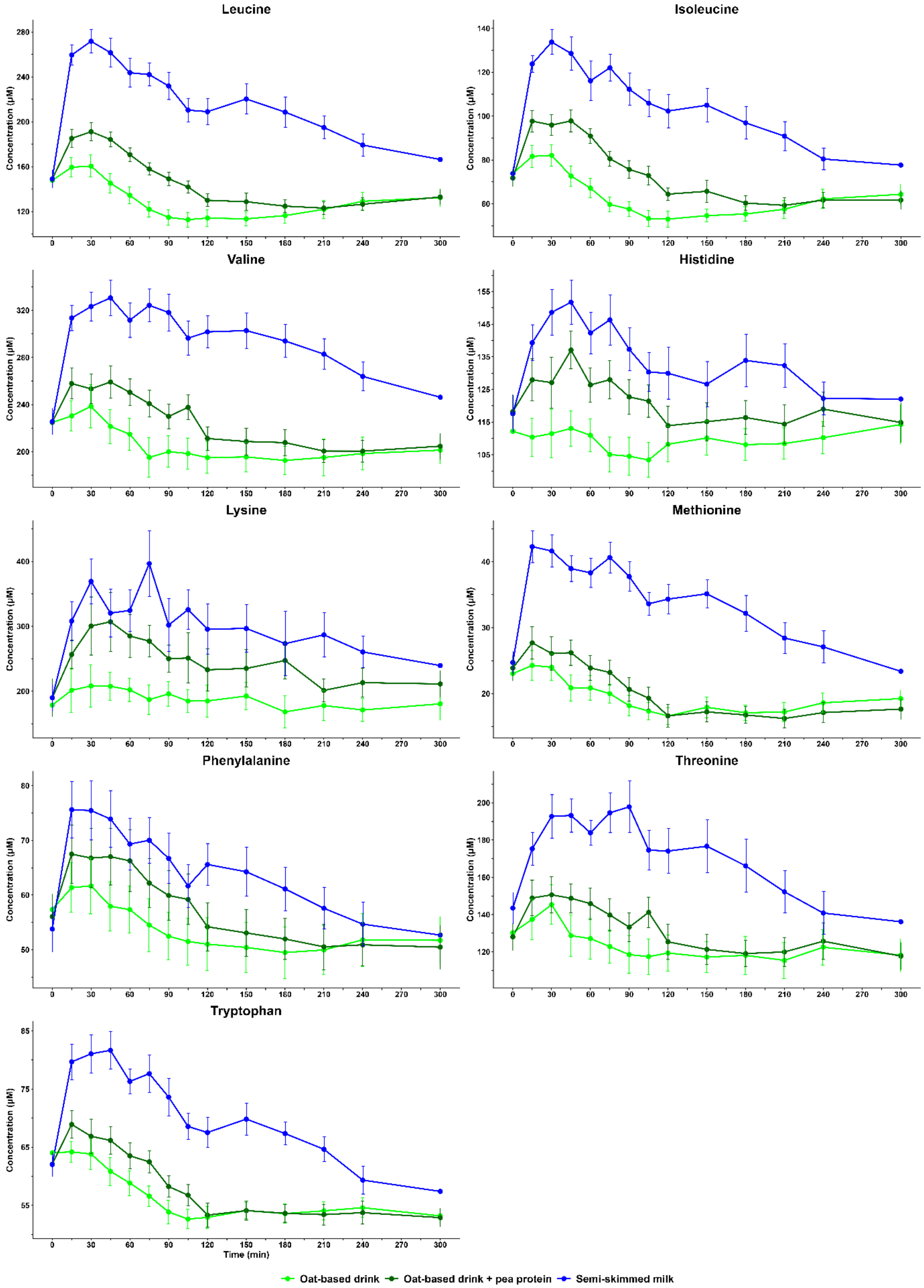


*Supplementary figure 3. Mean ± SE essential amino acid concentrations over time for the three drinks. Branched-chain amino acids are shown first, then other EAA's.*

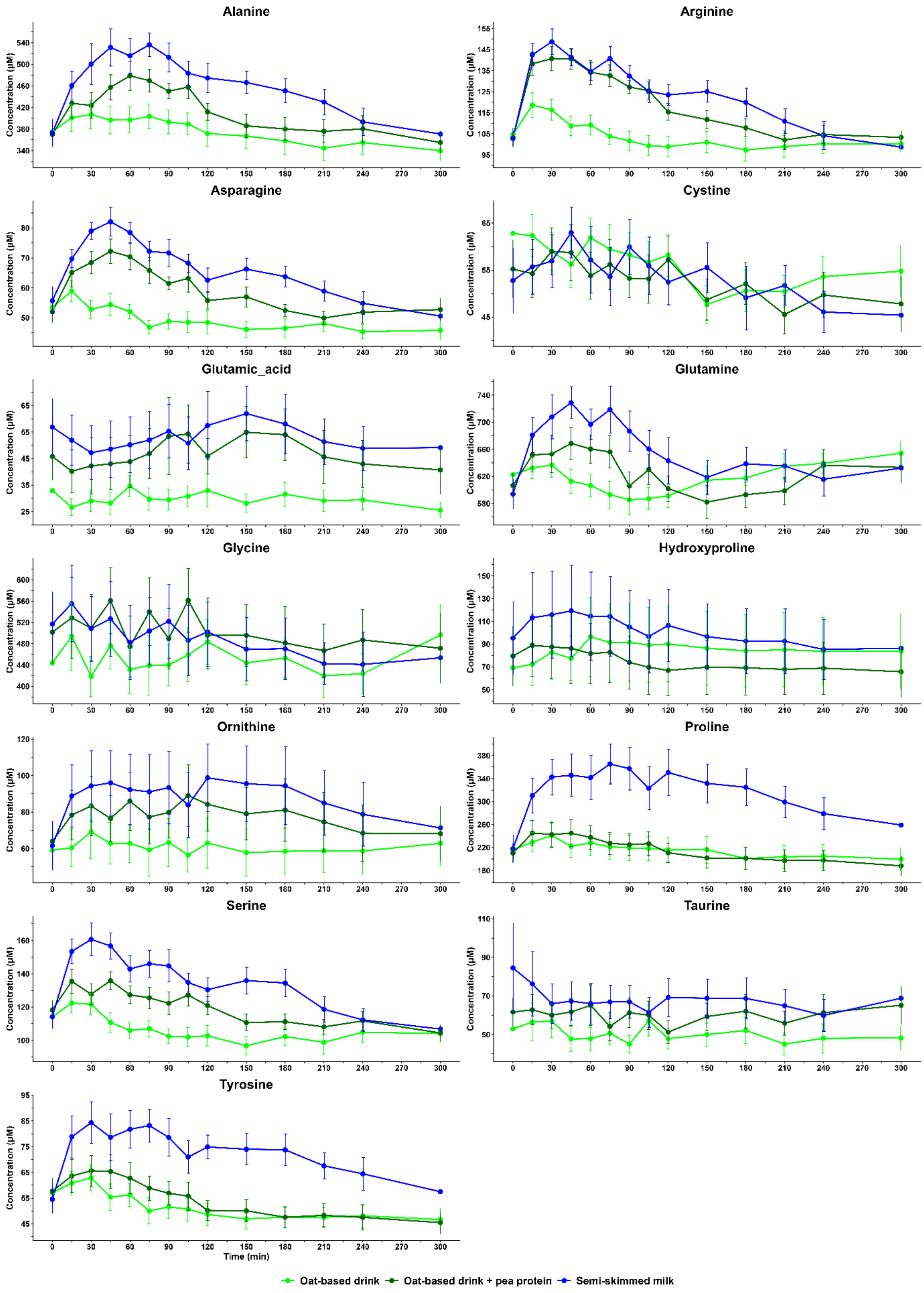


*Supplementary figure 4. Mean ± SE non-essential and conditionally essential amino acid concentrations over time for the three drinks.*

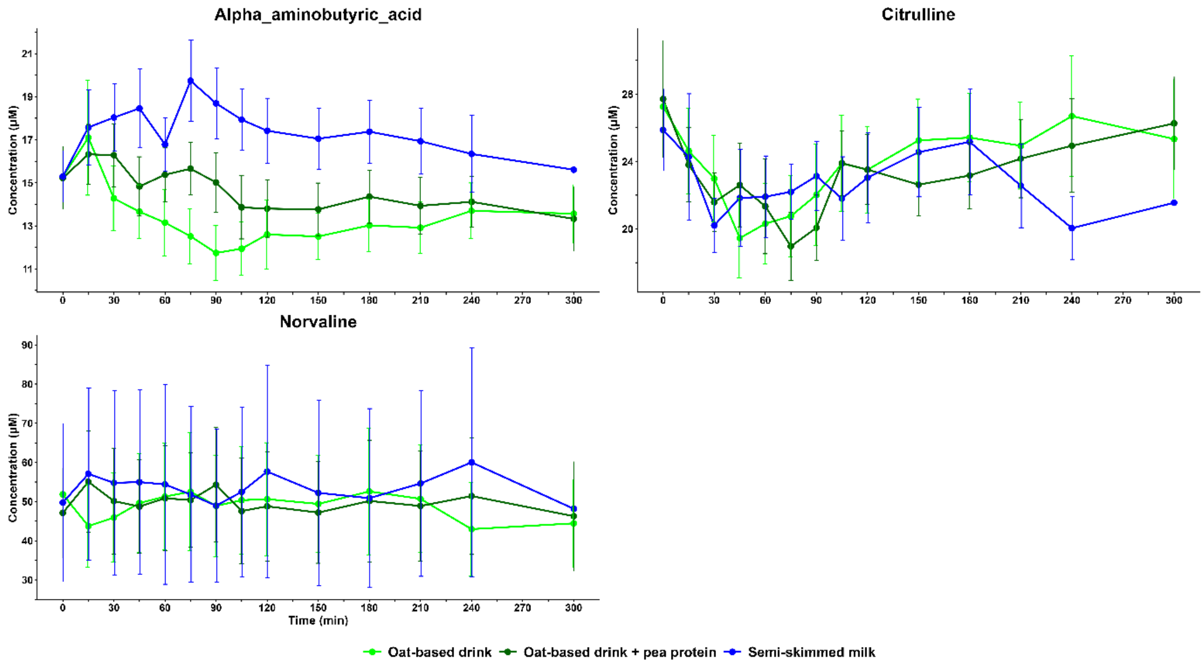


*Supplementary figure 5. Mean ± SE non-proteinogenic amino acid concentrations over time for the three drinks.*

**Supplementary table 4**. 5-h amino acid iAUC (µM*min) differences between the three drinks.

| Variable | Contrast | Mean difference in iAUC (95% CI) | P-value (tukey-corrected) |
|---|---|---|---|
| Total AA | OBD - (OBD+Pea) | -30154 (-82370, 22063) | 0.332 |
| | OBD - Milk | -169706 (-222549, -116862) | **<0.001** |
| | (OBD+Pea) - Milk | -139552 (-191776, -87328) | **<0.001** |
| EAA | OBD - (OBD+Pea) | -13549 (-35193, 8095) | 0.277 |
| | OBD - Milk | -91748 (-113447, -70049) | **<0.001** |
| | (OBD+Pea) - Milk | -78199 (-99854, -56544) | **<0.001** |
| | *Essential amino acids (BCAA first)* | | |
| Leucine | OBD - (OBD+Pea) | -1922 (-5123, 1279) | 0.305 |
| | OBD - Milk | -18358 (-21559, -15157) | **<0.001** |
| | (OBD+Pea) - Milk | -16436 (-19637, -13235) | **<0.001** |
| Isoleucine | OBD - (OBD+Pea) | -1597 (-3782, 587) | 0.180 |
| | OBD - Milk | -8107 (-10287, -5926) | **<0.001** |
| | (OBD+Pea) - Milk | -6509 (-8693, -4325) | **<0.001** |
| Valine | OBD - (OBD+Pea) | -1707 (-6311, 2896) | 0.625 |
| | OBD - Milk | -18832 (-23435, -14228) | **<0.001** |
| | (OBD+Pea) - Milk | -17124 (-21728, -12521) | **<0.001** |
| Histidine | OBD - (OBD+Pea) | -981 (-2717, 756) | 0.347 |

| **Variable** | **Contrast** | **Mean difference in iAUC (95% CI)** | **P-value (tukey-corrected)** |
|---|---|---|---|
| | OBD - Milk | -4212 (-5944, -2480) | **<0.001** |
| | (OBD+Pea) - Milk | -3231 (-4947, -1515) | **<0.001** |
| Lysine | OBD - (OBD+Pea) | -6942 (-21006, 7122) | 0.441 |
| | OBD - Milk | -23975 (-38039, -9910) | **<0.001** |
| | (OBD+Pea) - Milk | -17033 (-31070, -2996) | **0.016** |
| Methionine | OBD - (OBD+Pea) | -115 (-876, 646) | 0.923 |
| | OBD - Milk | -2698 (-3463, -1933) | **<0.001** |
| | (OBD+Pea) - Milk | -2583 (-3344, -1822) | **<0.001** |
| Phenylalanine | OBD - (OBD+Pea) | -777 (-2009, 455) | 0.272 |
| | OBD - Milk | -2791 (-4028, -1553) | **<0.001** |
| | (OBD+Pea) - Milk | -2014 (-3248, -780) | **0.001** |
| Threonine | OBD - (OBD+Pea) | -1321 (-3913, 1271) | 0.419 |
| | OBD - Milk | -8468 (-11125, -5812) | **<0.001** |
| | (OBD+Pea) - Milk | -7147 (-9827, -4467) | **<0.001** |
| Tryptophan | OBD - (OBD+Pea) | -204 (-1083, 676) | 0.830 |
| | OBD - Milk | -2235 (-3114, -1356) | **<0.001** |
| | (OBD+Pea) - Milk | -2031 (-2905, -1158) | **<0.001** |
| | *Non-essential and conditionally essential amino acids* | | |
| Alanine | OBD - (OBD+Pea) | -7287 (-15282, 709) | 0.078 |
| | OBD - Milk | -20308 (-28301, -12316) | **<0.001** |
| | (OBD+Pea) - Milk | -13021 (-21014, -5029) | **0.001** |
| Arginine | OBD - (OBD+Pea) | -3336 (-5192, -1479) | **<0.001** |
| | OBD - Milk | -4716 (-6573, -2858) | **<0.001** |
| | (OBD+Pea) - Milk | -1380 (-3235, 475) | 0.171 |
| Asparagine | OBD - (OBD+Pea) | -1808 (-2726, -891) | **<0.001** |
| | OBD - Milk | -3228 (-4147, -2310) | **<0.001** |
| | (OBD+Pea) - Milk | -1420 (-2346, -495) | **0.002** |
| Cystine | OBD - (OBD+Pea) | -140 (-1234, 954) | 0.945 |
| | OBD - Milk | -667 (-1770, 436) | 0.300 |
| | (OBD+Pea) - Milk | -527 (-1611, 556) | 0.451 |
| Glutamic acid | OBD - (OBD+Pea) | -2027 (-3928, -126) | **0.035** |
| | OBD - Milk | -1438 (-3442, 566) | 0.191 |
| | (OBD+Pea) - Milk | 589 (-1300, 2479) | 0.716 |
| Glutamine | OBD - (OBD+Pea) | -1039 (-10189, 8111) | 0.956 |
| | OBD - Milk | -11355 (-20675, -2034) | **0.015** |
| | (OBD+Pea) - Milk | -10315 (-19436, -1194) | **0.025** |
| Glycine | OBD - (OBD+Pea) | -1106 (-8127, 5915) | 0.917 |
| | OBD - Milk | 2212 (-4929, 9353) | 0.719 |
| | (OBD+Pea) - Milk | 3319 (-3509, 10146) | 0.452 |
| Hydroxyproline | OBD - (OBD+Pea) | 6284 (-4361, 16929) | 0.317 |
| | OBD - Milk | 2349 (-8459, 13157) | 0.849 |

| Variable | Contrast | Mean difference in iAUC (95% CI) | P-value (tukey-corrected) |
|---|---|---|---|
| | (OBD+Pea) - Milk | -3935 (-14620, 6749) | 0.629 |
| Ornithine | OBD - (OBD+Pea) | -2576 (-6193, 1041) | 0.195 |
| | OBD - Milk | -5750 (-9364, -2137) | **0.002** |
| | (OBD+Pea) - Milk | -3174 (-6787, 439) | **0.092** |
| Proline | OBD - (OBD+Pea) | 1024 (-7327, 9374) | 0.949 |
| | OBD - Milk | -24279 (-32624, -15935) | **<0.001** |
| | (OBD+Pea) - Milk | -25303 (-33664, -16943) | **<0.001** |
| Serine | OBD - (OBD+Pea) | -1490 (-3662, 682) | 0.218 |
| | OBD - Milk | -5532 (-7694, -3370) | **<0.001** |
| | (OBD+Pea) - Milk | -4042 (-6213, -1870) | **<0.001** |
| Taurine | OBD - (OBD+Pea) | -468 (-1985, 1049) | 0.721 |
| | OBD - Milk | -602 (-2202, 998) | 0.616 |
| | (OBD+Pea) - Milk | -134 (-1692, 1423) | 0.974 |
| Tyrosine | OBD - (OBD+Pea) | -336 (-2040, 1368) | 0.873 |
| | OBD - Milk | -5099 (-6806, -3392) | **<0.001** |
| | (OBD+Pea) - Milk | -4763 (-6472, -3053) | **<0.001** |
| | | *Non-proteinogenic amino acids* | |
| Alpha-aminobutyricacid | OBD - (OBD+Pea) | 105 (-520, 731) | 0.906 |
| | OBD - Milk | -497 (-1122, 129) | 0.137 |
| | (OBD+Pea) - Milk | -602 (-1228, 23) | 0.061 |
| Citrulline | OBD - (OBD+Pea) | 93 (-264, 449) | 0.792 |
| | OBD - Milk | 78 (-281, 436) | 0.850 |
| | (OBD+Pea) - Milk | -15 (-374, 344) | 0.994 |
| Norvaline | OBD - (OBD+Pea) | -1045 (-3544, 1454) | 0.552 |
| | OBD - Milk | -1846 (-4344, 651) | 0.174 |
| | (OBD+Pea) - Milk | -801 (-3299, 1696) | 0.702 |

Glucose and Insulin results tables

**Supplementary Table 5**. Treatment differences in estimated marginal mean glucose concentration (95%CI) per timepoint, adjusted for baseline glucose.

| **Timepoint (min)** | **Contrast** | **Mean difference in glucose concentration (95% CI)** | **P-value (tukey-corrected)** |
|---|---|---|---|
| T=15 | OBD - (OBD+Pea) | -0.09 (-0.63, 0.45) | 0.921 |
| | OBD - Milk | 0.86 (0.32, 1.40) | **<0.001** |
| | (OBD+Pea) - Milk | 0.95 (0.41, 1.49) | **<0.001** |
| T=30 | OBD - (OBD+Pea) | -0.06 (-0.59, 0.48) | 0.968 |
| | OBD - Milk | 1.68 (1.13, 2.22) | **<0.001** |
| | (OBD+Pea) - Milk | 1.73 (1.19, 2.27) | **<0.001** |
| T=45 | OBD - (OBD+Pea) | 0.20 (-0.33, 0.74) | 0.648 |
| | OBD - Milk | 1.22 (0.68, 1.76) | **<0.001** |
| | (OBD+Pea) - Milk | 1.02 (0.48, 1.55) | **<0.001** |
| T=60 | OBD - (OBD+Pea) | 0.14 (-0.40, 0.67) | 0.822 |
| | OBD - Milk | 0.37 (-0.17, 0.91) | 0.247 |
| | (OBD+Pea) - Milk | 0.23 (-0.31, 0.77) | 0.569 |
| T=75 | OBD - (OBD+Pea) | -0.12 (-0.67, 0.43) | 0.860 |
| | OBD - Milk | -0.06 (-0.61, 0.48) | 0.957 |
| | (OBD+Pea) - Milk | 0.06 (-0.49, 0.61) | 0.968 |
| T=90 | OBD - (OBD+Pea) | -0.21 (-0.75, 0.32) | 0.619 |
| | OBD - Milk | -0.31 (-0.86, 0.23) | 0.360 |
| | (OBD+Pea) - Milk | -0.10 (-0.64, 0.44) | 0.898 |
| T=105 | OBD - (OBD+Pea) | -0.07 (-0.61, 0.47) | 0.947 |
| | OBD - Milk | -0.07 (-0.62, 0.47) | 0.946 |
| | (OBD+Pea) - Milk | -0.00 (-0.54, 0.54) | 1.000 |
| T=120 | OBD - (OBD+Pea) | 0.01 (-0.53, 0.55) | 0.999 |
| | OBD - Milk | -0.06 (-0.61, 0.48) | 0.957 |
| | (OBD+Pea) - Milk | -0.08 (-0.62, 0.46) | 0.941 |
| T=150 | OBD - (OBD+Pea) | 0.14 (-0.40, 0.67) | 0.822 |
| | OBD - Milk | -0.16 (-0.70, 0.39) | 0.776 |
| | (OBD+Pea) - Milk | -0.29 (-0.83, 0.25) | 0.408 |
| T=180 | OBD - (OBD+Pea) | 0.16 (-0.38, 0.70) | 0.760 |
| | OBD - Milk | 0.11 (-0.43, 0.65) | 0.882 |
| | (OBD+Pea) - Milk | -0.05 (-0.59, 0.49) | 0.973 |
| T=210 | OBD - (OBD+Pea) | 0.36 (-0.18, 0.90) | 0.255 |
| | OBD - Milk | 0.19 (-0.36, 0.73) | 0.701 |
| | (OBD+Pea) - Milk | -0.18 (-0.72, 0.36) | 0.722 |
| T=240 | OBD - (OBD+Pea) | 0.07 (-0.47, 0.61) | 0.950 |
| | OBD - Milk | -0.02 (-0.57, 0.52) | 0.994 |
| | (OBD+Pea) - Milk | -0.09 (-0.63, 0.45) | 0.913 |
| T=300 | OBD - (OBD+Pea) | 0.12 (-0.42, 0.66) | 0.860 |

| | | |
|---|---|---|
| OBD - Milk | -0.02 (-0.57, 0.52) | 0.994 |
| (OBD+Pea) - Milk | -0.14 (-0.68, 0.40) | 0.807 |

**Supplementary Table 6**. Ratio of geometric mean insulin concentration (95%CI) per timepoint.

| **Timepoint (min)** | **Contrast** | **Ratio of geometric mean insulin concentration (95%CI)** | **P-value (tukey-corrected)** |
|---|---|---|---|
| T=15 | OBD - (OBD+Pea) | 1.13 (0.69, 1.85) | 0.834 |
| | OBD - Milk | 0.89 (0.55, 1.46) | 0.857 |
| | (OBD+Pea) - Milk | 0.79 (0.48, 1.30) | 0.511 |
| T=30 | OBD - (OBD+Pea) | 1.03 (0.63, 1.69) | 0.990 |
| | OBD - Milk | 1.89 (1.15, 3.09) | **0.007** |
| | (OBD+Pea) - Milk | 1.84 (1.12, 3.00) | **0.011** |
| T=45 | OBD - (OBD+Pea) | 0.92 (0.56, 1.51) | 0.921 |
| | OBD - Milk | 2.34 (1.43, 3.83) | **<0.001** |
| | (OBD+Pea) - Milk | 2.54 (1.55, 4.15) | **<0.001** |
| T=60 | OBD - (OBD+Pea) | 1.02 (0.62, 1.67) | 0.995 |
| | OBD - Milk | 2.14 (1.31, 3.50) | **<0.001** |
| | (OBD+Pea) - Milk | 2.09 (1.28, 3.43) | **0.001** |
| T=75 | OBD - (OBD+Pea) | 1.15 (0.69, 1.90) | 0.801 |
| | OBD - Milk | 1.46 (0.89, 2.39) | 0.167 |
| | (OBD+Pea) - Milk | 1.28 (0.77, 2.11) | 0.494 |
| T=90 | OBD - (OBD+Pea) | 0.98 (0.60, 1.60) | 0.993 |
| | OBD - Milk | 1.14 (0.69, 1.86) | 0.817 |
| | (OBD+Pea) - Milk | 1.16 (0.71, 1.90) | 0.753 |
| T=105 | OBD - (OBD+Pea) | 0.80 (0.49, 1.32) | 0.554 |
| | OBD - Milk | 0.92 (0.56, 1.51) | 0.926 |
| | (OBD+Pea) - Milk | 1.15 (0.70, 1.88) | 0.785 |
| T=120 | OBD - (OBD+Pea) | 0.99 (0.60, 1.61) | 0.997 |
| | OBD - Milk | 0.67 (0.41, 1.10) | 0.146 |
| | (OBD+Pea) - Milk | 0.68 (0.42, 1.12) | 0.169 |
| T=150 | OBD - (OBD+Pea) | 1.24 (0.76, 2.04) | 0.555 |
| | OBD - Milk | 0.71 (0.43, 1.16) | 0.224 |
| | (OBD+Pea) - Milk | 0.57 (0.35, 0.93) | **0.020** |
| T=180 | OBD - (OBD+Pea) | 1.17 (0.71, 1.91) | 0.740 |
| | OBD - Milk | 0.74 (0.45, 1.21) | 0.330 |
| | (OBD+Pea) - Milk | 0.64 (0.39, 1.04) | *0.079* |
| T=210 | OBD - (OBD+Pea) | 1.24 (0.76, 2.04) | 0.555 |
| | OBD - Milk | 0.86 (0.52, 1.40) | 0.744 |
| | (OBD+Pea) - Milk | 0.69 (0.42, 1.13) | 0.181 |
| T=240 | OBD - (OBD+Pea) | 1.27 (0.77, 2.08) | 0.493 |
| | OBD - Milk | 1.14 (0.69, 1.86) | 0.815 |
| | (OBD+Pea) - Milk | 0.90 (0.55, 1.47) | 0.858 |
| T=300 | OBD - (OBD+Pea) | 1.18 (0.72, 1.93) | 0.714 |
| | OBD - Milk | 1.14 (0.70, 1.87) | 0.801 |
| | (OBD+Pea) - Milk | 0.97 (0.59, 1.59) | 0.988 |

Gastric content: subvolume plots

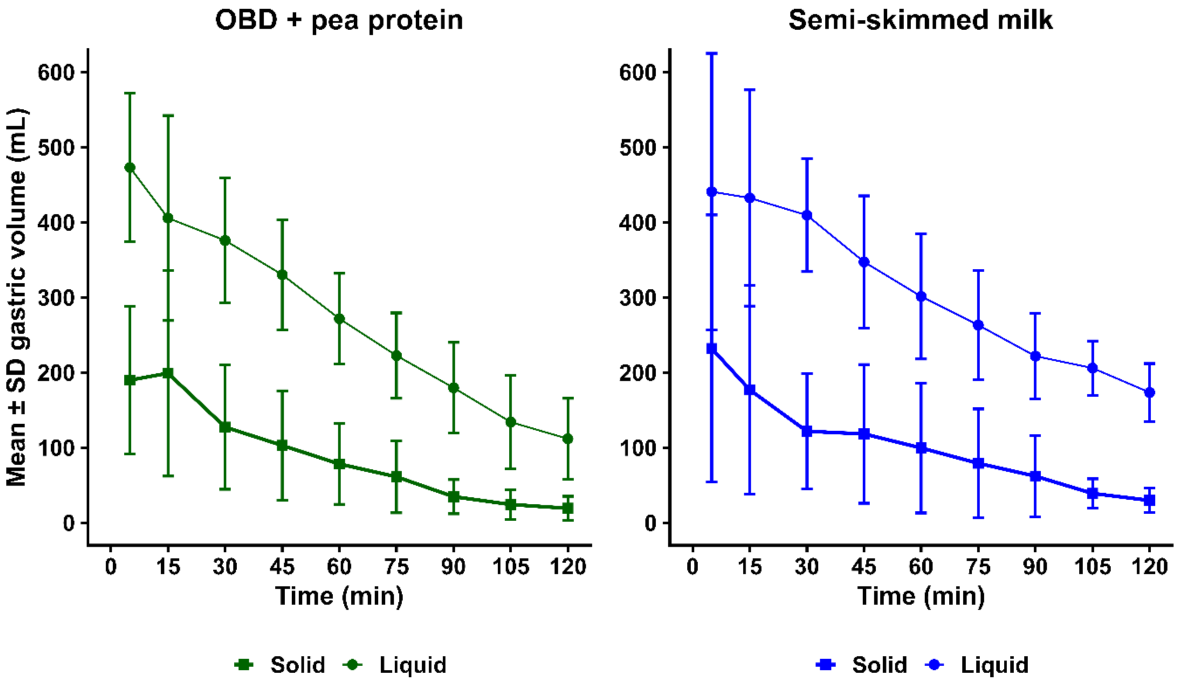


*Supplementary Figure 6. Mean ± SD gastric content subvolumes over time for the two MRI treatments. Solid indicates the summed volume of darker voxels and Liquid of brighter voxels at each timepoint, as classified with Otsu thresholding.*

Gastric content: image texture metric plots

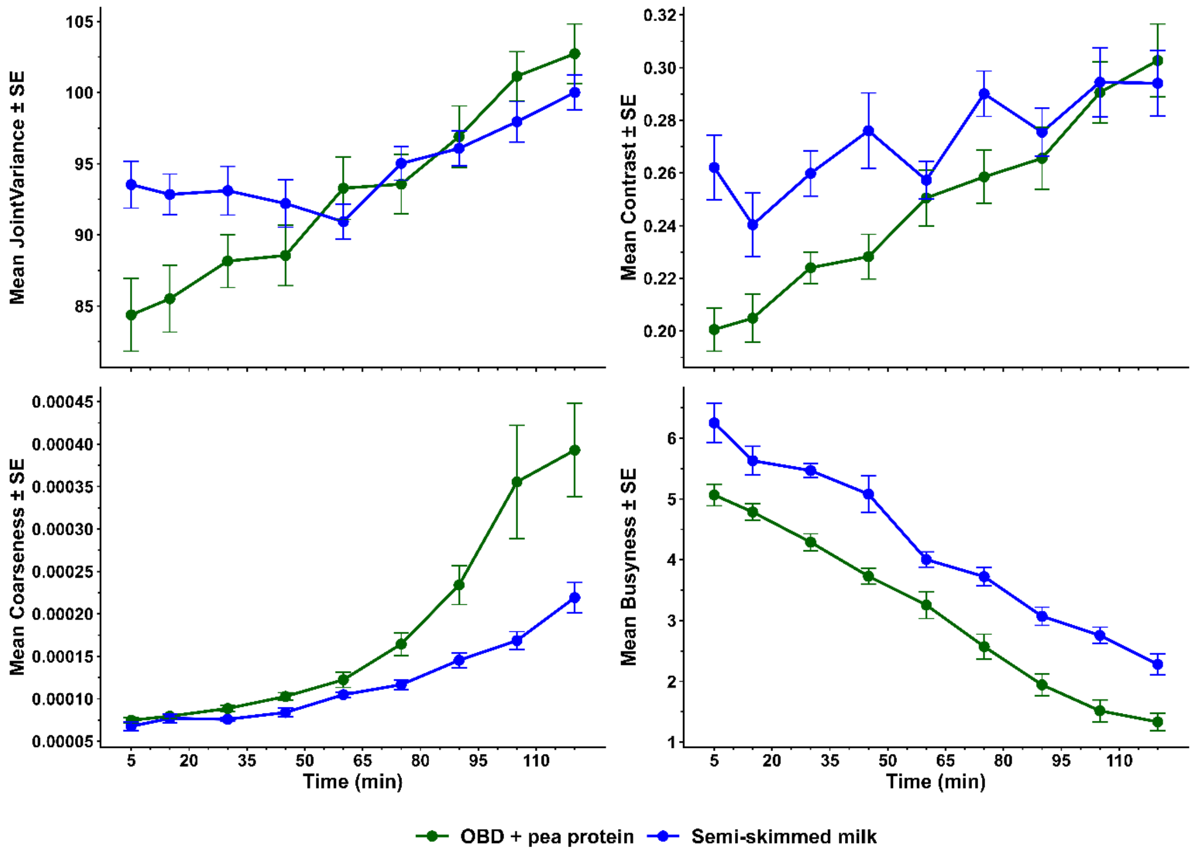


*Supplementary Figure 7. Mean ± SE postprandial gastric content image metrics over time for the two MRI treatments.*

FGF21 plot

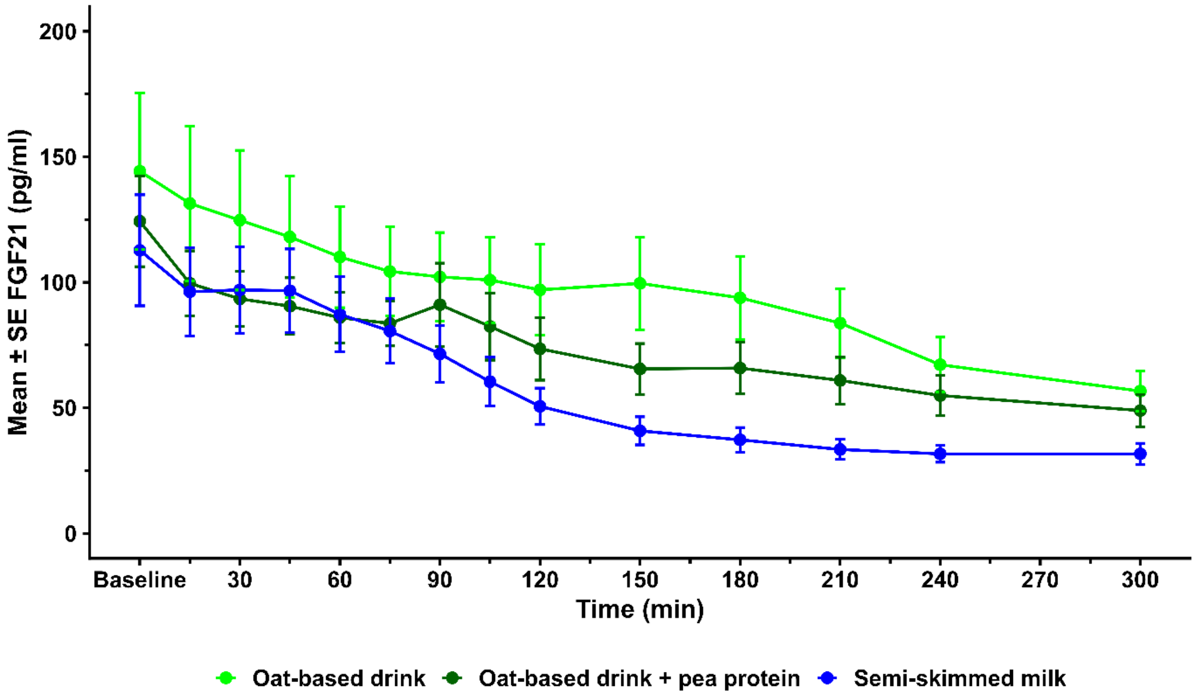


*Supplementary figure 8. Mean ±SE FGF21 concentration over time for each treatment.*

Subjective rating plots

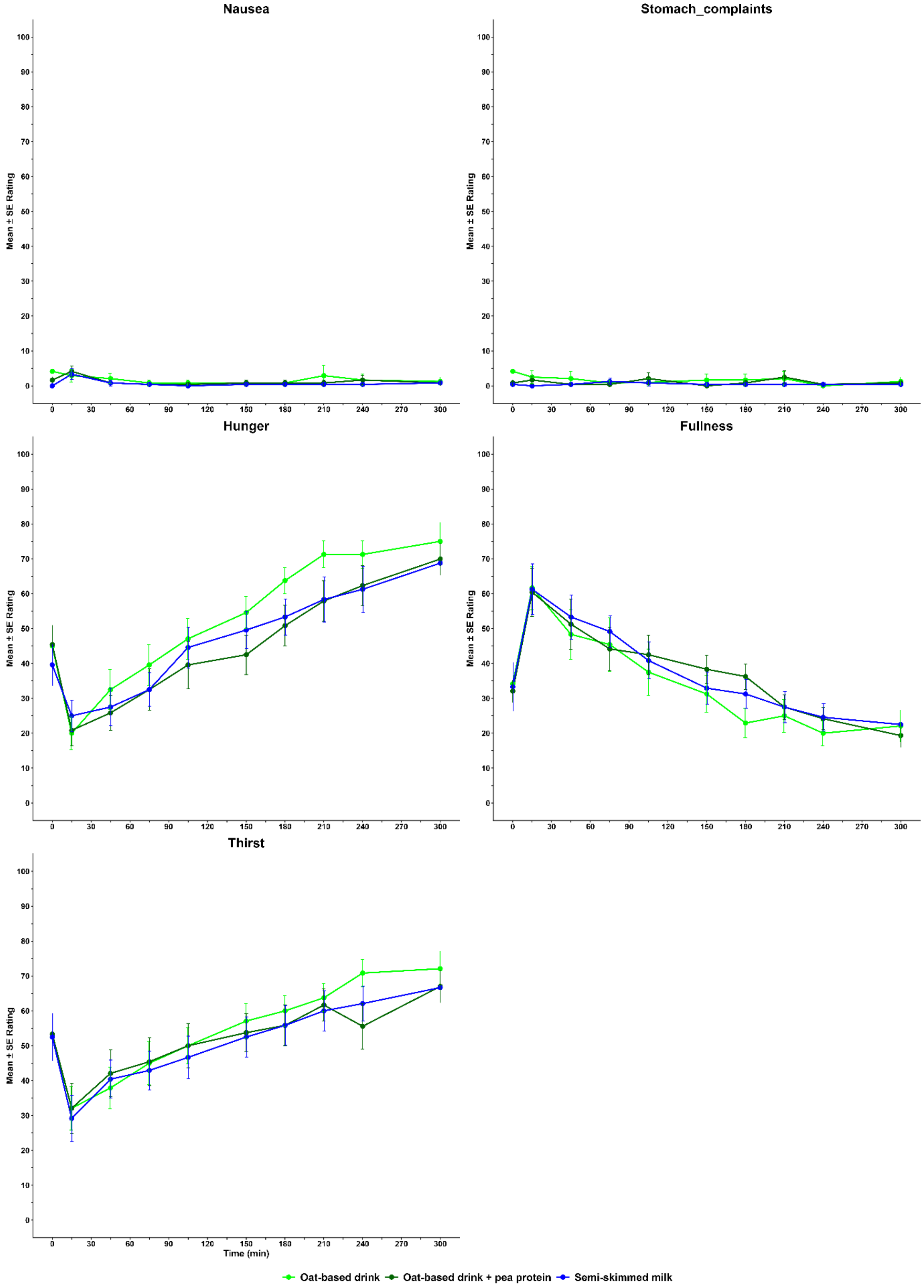


*Supplementary figure 9. Mean ± SE verbal subjective ratings (100-unit scale).*
